# Multimodal Artificial Intelligence for Cause-of-Death Assignment From Verbal Autopsy Records: A National Benchmark Study in South Africa

**DOI:** 10.64898/2026.10.15.26364476

**Authors:** Yue Chu, Zhenke Wu, Tyler H. McCormick, Zehang Richard Li, Samuel J. Clark

## Abstract

**Background:** Timely cause-of-death (COD) information guides health policy, yet many deaths in low- and middle-income countries are not medically certified. In Africa, fewer than 10% of deaths are. Verbal autopsy (VA) yields a structured symptom questionnaire, and a free-text narrative. Mortality surveillance increasingly records the narrative alongside the questionnaire, but the algorithms used to produce VA-based statistics were designed for the structured responses only, so narratives inform physician COD assignment but do not contribute to algorithmic COD assignment. We evaluated whether supervised models could use both modalities to improve COD assignment.

**Methods:** We analyzed 4,999 VA records for deaths at ages 12 years or older from a national sample of registered deaths in South Africa (RSA), using physician-assigned CODs as the reference standard. We trained unimodal and multimodal pipelines on the labelled records: pretrained language models (PLMs) fine-tuned on narratives alone, machine learning (ML) models on structured responses alone, and the two combined at the data, feature and decision levels. We also prompted a locally deployed generative large language model (LLM) on the same unimodal and multimodal inputs, without parameter updates. All were compared with InSilicoVA, the operational comparator representing current automated practice. We evaluated individual-level assignment, population-level agreement, calibration, and generalization across provinces in RSA, reporting these metrics rather than ranking pipelines on any one of them.

**Results:** Every supervised pipeline improved on InSilicoVA by 8.5–15.9 percentage points in top-1 accuracy and 3.5–7.1 in cause-specific mortality fraction (CSMF) accuracy. The narrative and structured pipelines performed similarly in aggregate but differed by cause: 63.9% of deaths received identical predictions, 11.6% were correct only from the narrative pipeline and 13.6% only from the questionnaire pipeline, with the narrative pipeline more accurate for noncommunicable causes and the questionnaire pipeline for communicable causes. Against the unimodal approaches, the decision-fusion pipelines added 4.2 to 5.4 percentage points in top-1 accuracy, concentrated in the most common causes, whereas feature fusion with cross-modal attention improved the less common causes instead, adding 6.6 (1.7–11.5) points in balanced accuracy and 5.5 (0.6–10.3) in macro F1. Feature fusion reached the highest CSMF accuracy (95.7%). Paired differences from the better unimodal representative ranged from *−*0.2 to +3.4 points, with all intervals including zero. The LLM exceeded InSilicoVA on individual-level metrics and reached the highest balanced accuracy of any pipeline (60.6%), while the supervised pipelines retained higher CSMF accuracy. Supervised pipelines had lower expected calibration error than InSilicoVA. Supervised gains held across all nine held-out provinces, including every top-1 comparison and nearly every macro F1 and CSMF comparison.

**Conclusions:** PLMs using narratives alone substantially improved on the current best practice and matched ML pipelines using structured responses, while multiple multimodal strategies produced further gains in individual COD assignment. The zero-shot LLM showed particular strength on rare causes, whereas supervised pipelines better recovered the population cause distribution. These findings support incorporating narratives as a core input to automated VA systems and evaluating representative fusion strategies for the intended output. External validation across settings and age groups will support broader implementation.

## 1 Introduction

Cause-of-death (COD) information guides burden of disease estimates, public health interventions, and resource allocation, yet many of the approximately 55 million deaths worldwide each year are not registered with a usable medical cause, especially in low- and middle-income countries (LMICs). In Africa, fewer than 10% of deaths have a medically certified COD.^1^ In these settings, verbal autopsy (VA) is the principal tool for assigning causes at population scale. Trained fieldworkers interview a caregiver or next of kin about the period before death, producing an inherently multimodal record: a structured questionnaire of discrete symptom responses and a free-text narrative describing medical history, illness progression, care-seeking, and circumstances of death.^2–4^ Physician review ascertains COD using both components but is slow, costly, and difficult to reproduce or scale. The automated algorithms that produce VA-based mortality statistics, such as InSilicoVA and InterVA, are inexpensive, reproducible, and scalable, but were designed before modern natural-language processing (NLP) and use only the structured responses.^5,6^ Where narratives are collected, the information they contain is therefore available to physicians but discarded by the methods used at scale.

Computational work on VA narratives has established COD-relevant signal across engineered linguistic features, embeddings, neural networks, and domain-adapted or contextual representations, largely using the narrative as the sole input.^7–12^ Recent studies have applied GPT models to elicited narratives^13^ or text reconstructed from structured responses,^14^ or used narrative-only predictions to improve downstream population inference.^15^ Multimodal studies have reported cause-specific gains from concatenation,^7,16^ improvements from jointly learned narrative and questionnaire representations,^17^ and more modest gains from decision-level stacking.^18^ LAVA evaluated joint-input language and embedding models, a structured latent-class model, and their ensembles, but did not include matched narrative-only and structured-only ablations of the joint-input models.^19^ Because these studies differed in datasets, inputs, cause lists, and fusion designs, the relative contributions of each modality and fusion level—particularly by cause and for population estimates—have not been compared within a common benchmark. Answering these questions requires evaluating narrative-only, structured-only, and fused models on the same records, labels, and partitions against the algorithm used in practice (Supplementary Section S1).

Using VA records from a national validation study in South Africa, with reference causes assigned by physicians who reviewed the complete record, we compared narrative-based pretrained language models (PLMs; general-domain and domain-adapted encoders), structured-input machine learning (ML), and fusion of the two at the data, feature, and decision levels — combining inputs, learned representations, or predictions — benchmarking all approaches against InSilicoVA^5^ as current best practice in automated VA-based COD assignment. In addition to the supervised pipelines, we evaluated an unsupervised approach that assigns causes without fitting any parameters to the training labels, prompting a generative large language model (LLM) on the same inputs. We asked whether supervised models improve on that practice, whether the narrative and the questionnaire contribute differently, whether the level at which they are combined matters, and how unsupervised models perform. We also assessed agreement with the population cause distribution, calibration, error patterns, and stability across provinces and analytic choices.

## 2 Methods

### 2.1 Data sources and study population

We used VA records from the South African National Cause-of-Death Validation Project (SANCODV), which was designed as a nationally representative sample of registered deaths drawn for cause-of-death validation.^20^ SANCODV conducted VA interviews for deaths registered from September 2017 through April 2018 in 27 subdistricts selected at random across South Africa, with pseudo-stratification by socioeconomic status. Fieldworkers were thoroughly trained and administered the WHO 2016 standardized VA instrument to a close caregiver or next-of-kin, and narratives were transcribed into English. The reference COD was assigned by 75 trained physicians who independently reviewed the full record, narrative and questionnaire together, under a parallel review process with adjudication, and coded causes to the International Classification of Diseases, 10th Revision (ICD-10).^21^ Unlike VA datasets assembled within health and demographic surveillance sites, SANCODV draws on registered deaths from all nine provinces, and every analyzed record carries both the narrative and the questionnaire together with a physician-assigned reference cause.^20^

We included 4,999 VAs for deaths at ages 12 years and older with valid narratives. Structured features were harmonized to the indicator set expected by InSilicoVA,^22^ and narratives underwent conservative copy-editing with a large language model, limited to spelling, punctuation, grammar, and same-concept shorthand standardization. Physician-coded ICD-10 assignments were grouped into the 19 COD categories used for training and evaluation, which retain the causes of greatest public-health interest while limiting sparsity. There were at least 24 deaths per modeled cause. Preprocessing, quality assurance, and the grouping schema are in Supplementary Sections S2–S3 and Table S1.

### 2.2 Modeling strategies

VAs were randomly split into an 80% training set and a 20% held-out test set, stratified by COD group. InSilicoVA, run through the openVA package with cause-specific symptom probabilities estimated from the training data, served as the operational comparator. Its symptom-given-cause probabilities were grouped into ranked levels by fixed cut-points, with the level values reestimated during sampling.^5,22^

#### Unimodal strategies

We fine-tuned general-domain and domain-adapted encoders end-to-end on only the narratives: BERT, BlueBERT, BioClinicalBERT, ClinicalBERT, BiomedBERT, and BioMed-RoBERTa.^23–27^ Structured-question models used the harmonized structured indicators with supervised ML classifiers: LightGBM, XGBoost, CatBoost, gradient-boosted decision trees (GBDT), random forests, and multilayer perceptrons (MLP).^28–32^

#### Multimodal fusion strategies

*Data fusion* rendered the structured responses as text and appended them to the narrative, so that a single long-context PLM read the whole record.^33^ *Feature fusion* used AutoGluon AutoMM^34,35^ to encode the narrative and the structured responses separately and learn a joint representation. BioMed-RoBERTa was used as the narrative backbone encoding input tokens into dense vector embeddings for subsequent fusion. *Decision fusion* kept the pipelines independent and combined their predicted cause probabilities, by equal-weight soft voting, class-aware voting weighted by each model’s out-of-fold per-cause F1, or stacking with a second-stage meta-learner. All three combined the same ten base models, five narrative PLMs and five structured ML models. Soft voting averaged the probabilities produced by the unimodal pipelines themselves, whereas class-aware voting and stacking learned their combination rules from out-of-fold predictions obtained by refitting those ten models within the training partition. Base models, architectures, and checkpoints are in Supplementary Section S4.

#### Generative large language model

We additionally evaluated an unsupervised approach, in which an open-weight generative LLM, served locally through Ollama, assigned a cause from the same 19-cause list zero-shot, with no parameter updates. Here *unsupervised* denotes that no study labels were used for parameter fitting. Three arms read the same record content as the narrative-only, structured-only and data-fusion pipelines: the narrative alone, the structured responses rendered as text, and both together. The model, gpt-oss-120b, and the prompt configuration for each arm were chosen by macro F1 on a held-in subset of the training records, as for hyperparameter optimization above. Model selection and prompt configurations are in Supplementary Section S4.4.

Most pipeline families include more than one candidate model. The main text tables and figures present one representative per family, chosen on mean macro F1 in cross-validation confined to the training partition rather than on held-out performance. For feature fusion the architecture was specified in advance. All candidates and their held-out performance are reported in Supplementary Table S8.

### 2.3 Hyperparameter optimization

Hyperparameter optimization used Optuna with the Tree-structured Parzen Estimator sampler. Trials were run on the training set only, and the configuration with the highest mean macro F1 in 3-fold cross-validation was refit on the full training set before held-out evaluation. InSilicoVA was configured following the default setting used in published analyses.^22^ Sensitivity analyses of alternative InSilicoVA configurations are in Supplementary Section S4.3.1.

### 2.4 Performance evaluation

On the fixed held-out test set, individual-level metrics were top-1 accuracy, top-3 accuracy, balanced accuracy, macro F1, and weighted F1. Population-level agreement was measured with cause-specific mortality fraction (CSMF) accuracy and chance-corrected CSMF (CCCSMF) accuracy, using cause fractions computed from each pipeline’s top-1 assignments.^36^ These answer different questions, namely how often the assigned cause is right, how accurate the assignment is with every cause weighted equally, and how closely the predicted cause distribution matches the reference, so we report them alongside one another rather than ranking them. Calibration was assessed from each pipeline’s full probability matrix, using classwise expected calibration error (ECE), maximum calibration error (MCE), and the Brier score. Uncertainty was quantified by a nonparametric bootstrap (1,000 resamples of the test cases, with the 2.5th and 97.5th percentiles as 95% confidence intervals, CI). Comparisons between pipelines used a paired bootstrap of the differences. Metric definitions are in Supplementary Section S4.5.

### 2.5 Additional analyses

We examined the synergy between the narrative and structured questions by partitioning the held-out deaths according to which of the two unimodal representatives assigned the correct cause: both, one only, or neither, and how often the fusion model was correct within each group.

Two sensitivity analyses examined robustness to analytic and design choices: the granularity of the COD grouping; alternative feature-fusion backbone encoders and fusion strategies. We decomposed held-out accuracy into modality contributions with Shapley values, for the soft-voting ensemble over its ten member models and for the LLM over its narrative and structured inputs, overall and within communicable and noncommunicable reference causes. We also conducted a leave-one-province-out (LOPO) analysis, in which models were refit on eight provinces and evaluated on the ninth, with hyperparameters fixed at their globally selected values. Details are in Supplementary Sections S7 and S8.

## 3 Results

### 3.1 Dataset characteristics

Demographic characteristics are presented in Table 1. The underlying COD distribution was highly imbalanced, with HIV/AIDS the leading COD (23.2%) and indeterminate cause accounting for 15.6% of deaths (Supplementary Table S1). Narratives had a median length of 95 words (IQR 69–128; Supplementary Figure S1), well below the maximum input length of the encoders.

**Table 1:** Dataset and task characteristics. Per-cause CSMF distributions are in Supplementary Table S1.

| Characteristic | SANCODV |
| --- | --- |
| Study setting | National sample of registered deaths, South Africa |
| Total sample size | 4,999 |
| Sample size in held-out test | 1000 |
| Active causes in held-out test | 19 |
| Deaths due to indeterminate cause (%) | 15.6 |
| Narrative word count, median (IQR) | 95 (69–128) |
| Female decedents, n (%)<sup>*</sup> | 2,388 (47.8) |
| Age group, n (%)<sup>*</sup> |  |
| 65+ years | 1,682 (33.6) |
| 50–64 years | 1,324 (26.5) |
| 15–49 years | 1,966 (39.3) |
| 12–14 years | 27 (0.5) |
| Province, n (%) |  |
| Eastern Cape | 556 (11.1) |
| Free State | 617 (12.3) |
| Gauteng | 427 (8.5) |
| KwaZulu-Natal | 841 (16.8) |
| Limpopo | 309 (6.2) |
| Mpumalanga | 567 (11.3) |
| Northern Cape | 471 (9.4) |
| North West | 1,026 (20.5) |
| Western Cape | 185 (3.7) |
| Leading causes of deaths, n (%) |  |
| HIV/AIDS | 1,158 (23.2) |
| Indeterminate | 782 (15.6) |
| Cardiac disease | 515 (10.3) |
| Neoplasms | 445 (8.9) |
| Pulmonary tuberculosis | 311 (6.2) |

<sup>\*</sup> Gender and age of the decedent reported by survey respondent, usually close caregiver or next-of-kin of the decedent.

### 3.2 Performance of AI models across modalities

Table 2 reports the performance of the representative model from each pipeline, selected on cross-validated macro F1 within the training partition. The full leaderboard for all models is in Supplementary Table S8.

**Table 2:**
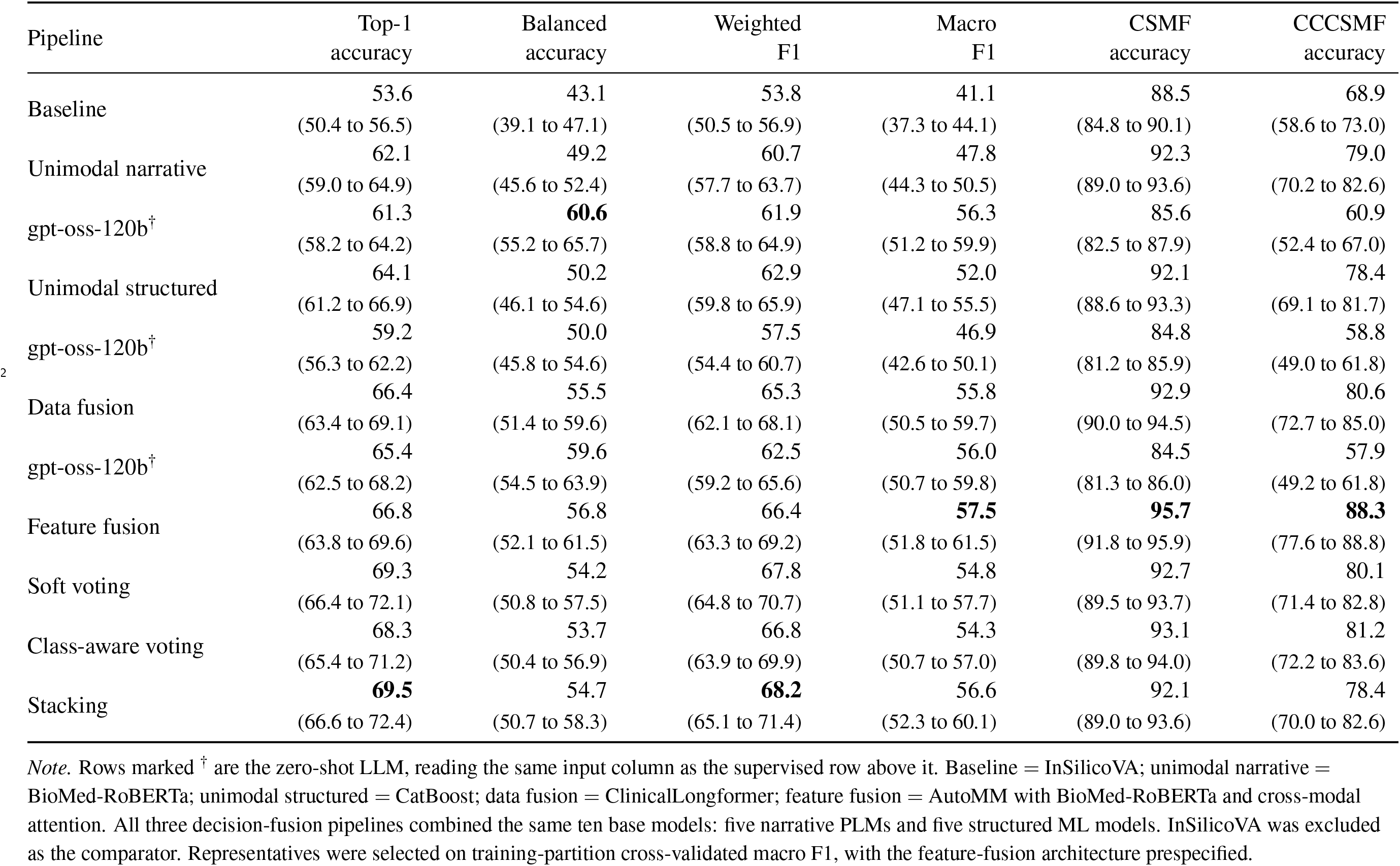
Model performance by pipeline. Representative pipelines are shown with 95% bootstrap confidence intervals (1,000 resamples of the held-out set). Bold marks the highest value within a metric.

| Pipeline | Top-1<br>accuracy | Balanced<br>accuracy | Weighted<br>F1 | Macro<br>F1 | CSMF<br>accuracy | CCCSMF<br>accuracy |
| --- | --- | --- | --- | --- | --- | --- |
| Baseline | 53.6<br>(50.4 to 56.5) | 43.1<br>(39.1 to 47.1) | 53.8<br>(50.5 to 56.9) | 41.1<br>(37.3 to 44.1) | 88.5<br>(84.8 to 90.1) | 68.9<br>(58.6 to 73.0) |
| Unimodal narrative | 62.1<br>(59.0 to 64.9) | 49.2<br>(45.6 to 52.4) | 60.7<br>(57.7 to 63.7) | 47.8<br>(44.3 to 50.5) | 92.3<br>(89.0 to 93.6) | 79.0<br>(70.2 to 82.6) |
| gpt-oss-120b<sup>†</sup> | 61.3<br>(58.2 to 64.2) | **60.6**<br>(55.2 to 65.7) | 61.9<br>(58.8 to 64.9) | 56.3<br>(51.2 to 59.9) | 85.6<br>(82.5 to 87.9) | 60.9<br>(52.4 to 67.0) |
| Unimodal structured | 64.1<br>(61.2 to 66.9) | 50.2<br>(46.1 to 54.6) | 62.9<br>(59.8 to 65.9) | 52.0<br>(47.1 to 55.5) | 92.1<br>(88.6 to 93.3) | 78.4<br>(69.1 to 81.7) |
| gpt-oss-120b<sup>†</sup> | 59.2<br>(56.3 to 62.2) | 50.0<br>(45.8 to 54.6) | 57.5<br>(54.4 to 60.7) | 46.9<br>(42.6 to 50.1) | 84.8<br>(81.2 to 85.9) | 58.8<br>(49.0 to 61.8) |
| Data fusion | 66.4<br>(63.4 to 69.1) | 55.5<br>(51.4 to 59.6) | 65.3<br>(62.1 to 68.1) | 55.8<br>(50.5 to 59.7) | 92.9<br>(90.0 to 94.5) | 80.6<br>(72.7 to 85.0) |
| gpt-oss-120b<sup>†</sup> | 65.4<br>(62.5 to 68.2) | 59.6<br>(54.5 to 63.9) | 62.5<br>(59.2 to 65.6) | 56.0<br>(50.7 to 59.8) | 84.5<br>(81.3 to 86.0) | 57.9<br>(49.2 to 61.8) |
| Feature fusion | 66.8<br>(63.8 to 69.6) | 56.8<br>(52.1 to 61.5) | 66.4<br>(63.3 to 69.2) | **57.5**<br>(51.8 to 61.5) | **95.7**<br>(91.8 to 95.9) | **88.3**<br>(77.6 to 88.8) |
| Soft voting | 69.3<br>(66.4 to 72.1) | 54.2<br>(50.8 to 57.5) | 67.8<br>(64.8 to 70.7) | 54.8<br>(51.1 to 57.7) | 92.7<br>(89.5 to 93.7) | 80.1<br>(71.4 to 82.8) |
| Class-aware voting | 68.3<br>(65.4 to 71.2) | 53.7<br>(50.4 to 56.9) | 66.8<br>(63.9 to 69.9) | 54.3<br>(50.7 to 57.0) | 93.1<br>(89.8 to 94.0) | 81.2<br>(72.2 to 83.6) |
| Stacking | **69.5**<br>(66.6 to 72.4) | 54.7<br>(50.7 to 58.3) | **68.2**<br>(65.1 to 71.4) | 56.6<br>(52.3 to 60.1) | 92.1<br>(89.0 to 93.6) | 78.4<br>(70.0 to 82.6) |

*Note.* Rows marked <sup>†</sup> are the zero-shot LLM, reading the same input column as the supervised row above it. Baseline = InSilicoVA; unimodal narrative = BioMed-RoBERTa; unimodal structured = CatBoost; data fusion = ClinicalLongformer; feature fusion = AutoMM with BioMed-RoBERTa and cross-modal attention. All three decision-fusion pipelines combined the same ten base models: five narrative PLMs and five structured ML models. InSilicoVA was excluded as the comparator. Representatives were selected on training-partition cross-validated macro F1, with the feature-fusion architecture prespecified.

The representative narrative PLM (BioMed-RoBERTa) reached 62.1% top-1 accuracy and 92.3% CSMF accuracy, and the representative structured model (CatBoost) 64.1% and 92.1%, against 53.6% and 88.5% for InSilicoVA: gains of 8.5 to 10.5 percentage points in top-1 accuracy and 3.5 to 3.7 points in CSMF accuracy. Each modality therefore carried substantial diagnostic information on its own.

The two modalities were close on top-1 accuracy and on agreement with the population cause distribution, and differed more in macro F1 (47.8% vs. 52.0%). Both exceeded the comparator on every metric, by larger margins on the individual-level metrics than on CSMF accuracy, where CatBoost’s margin was not distinguishable from zero (3.5 points, 95% CI *−*0.1 to 7.4).

The structured model and InSilicoVA used the same questionnaire indicators, yet differed by 10.5 percentage points in top-1 accuracy and 3.6 points in CSMF accuracy, and the difference was broad across causes: the structured model was more accurate for 13 of the 19 causes and less accurate for three, the largest difference being for pneumonia (Figure 1).

**Figure 1:**
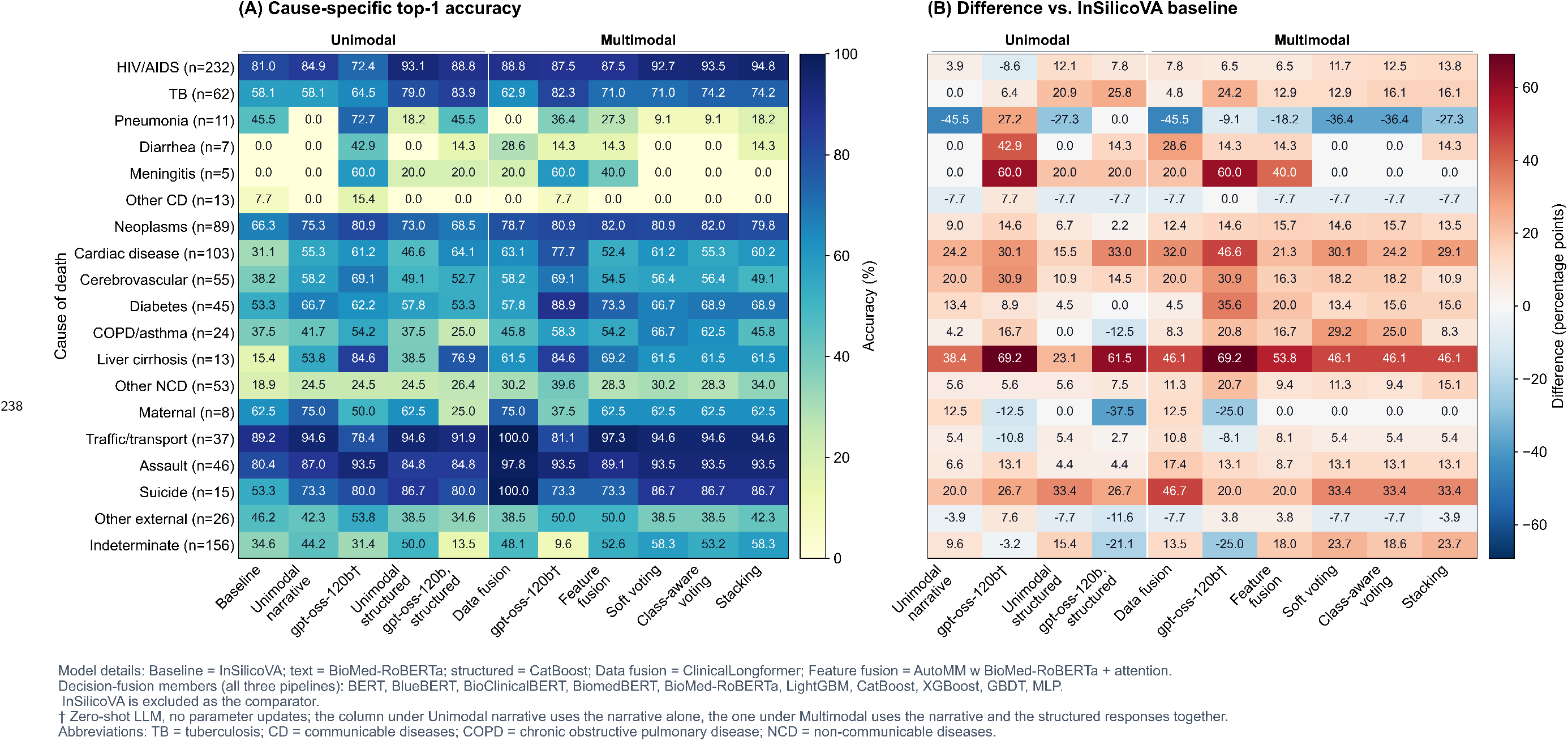
Cause-specific top-1 accuracy in SANCODV. Left: accuracy by pipeline. Right: percentage-point difference from InSilicoVA. Columns marked ^†^ are the zero-shot LLM, placed under the modality whose input column each one reads.

Multimodal fusion produced further gains with distinct strengths across strategies. Stacking had the highest top-1 accuracy (69.5%), whereas feature fusion with cross-modal attention was strongest on class-balanced metrics and CSMF accuracy, raising macro F1 from 52.0% for the better unimodal representative to 57.5%, and CSMF accuracy from 92.3% to 95.7%.

Relative to the better unimodal representative for each metric, the three decision-fusion pipelines gave the clearest gains in overall accuracy: stacking added 5.4 (3.1 to 7.7) percentage points in top-1 accuracy, soft voting 5.2 (2.9 to 7.6), and class-aware voting 4.2 (2.0 to 6.5). Their gains in balanced accuracy and macro F1 were smaller, and for soft voting and class-aware voting the intervals spanned zero. Stacking was the exception, at 4.4 (0.3 to 8.5) in balanced accuracy and 4.6 (0.5 to 8.9) in macro F1. Data-level and feature-level fusion showed the converse pattern: their top-1 gains spanned zero, whereas their balanced-accuracy gains did not, at 5.3 (0.3 to 10.5) and 6.6 (1.7 to 11.5), and feature fusion added 5.5 (0.6 to 10.3) in macro F1. Paired differences in CSMF accuracy from the better unimodal representative ranged from *−*0.2 to +3.4 points, with all intervals including zero.

The zero-shot LLM — gpt-oss-120b — exceeded InSilicoVA on all individual-level metrics from each of the three inputs (top-1 accuracy +7.7, +5.6 and +11.8 percentage points from the narrative, the structured responses and both together, balanced accuracy +17.5, +6.8 and +16.5, macro F1 +15.1, +5.7 and +14.8) and reached the highest balanced accuracy of any pipeline (60.6% with the narrative alone), with the improvement concentrated in the rarer causes (Figure 1), but it was the weakest on CSMF accuracy (2.9 to 4.0 points below InSilicoVA; Table 2, Supplementary Section S4.4). These family-level differences were seen for all candidates in each family (Supplementary Section S5).

### 3.3 Cause-specific performance

Every paired difference between the narrative-only and structured-only pipelines spanned zero: top-1 accuracy (*−*2.0 points [*−*5.1 to +1.0]), macro F1 (*−*4.1 [*−*8.5 to +0.7]), and CSMF accuracy (+0.2 [*−*2.8 to +3.4]). Despite aggregate similarity, we observed substantial between-cause heterogeneity (Figure 1). BioMed-RoBERTa was more accurate for noncommunicable diseases (NCDs) (aggregate accuracy 56.5% vs. 50.5%), with the largest advantages for liver cirrhosis and cerebrovascular disease. CatBoost was more accurate for communicable causes (81.2% vs. 70.6%), with the largest differences for pulmonary tuberculosis (TB) and pneumonia.

Decomposing the accuracy of the combined pipelines revealed a similar pattern of modality contributions in both the soft-voting ensemble and the LLM: the structured input contributed more within communicable causes and the narrative within noncommunicable causes (Figure 2A; Supplementary Section S7.2). Varying the modality composition of the soft-voting ensemble at fixed size showed that ensembling narrative and structured models together performed better at the individual level than ensembling models of a single modality. Population-level performance varied little across modality compositions (Supplementary Section S7.2).

**Figure 2:**
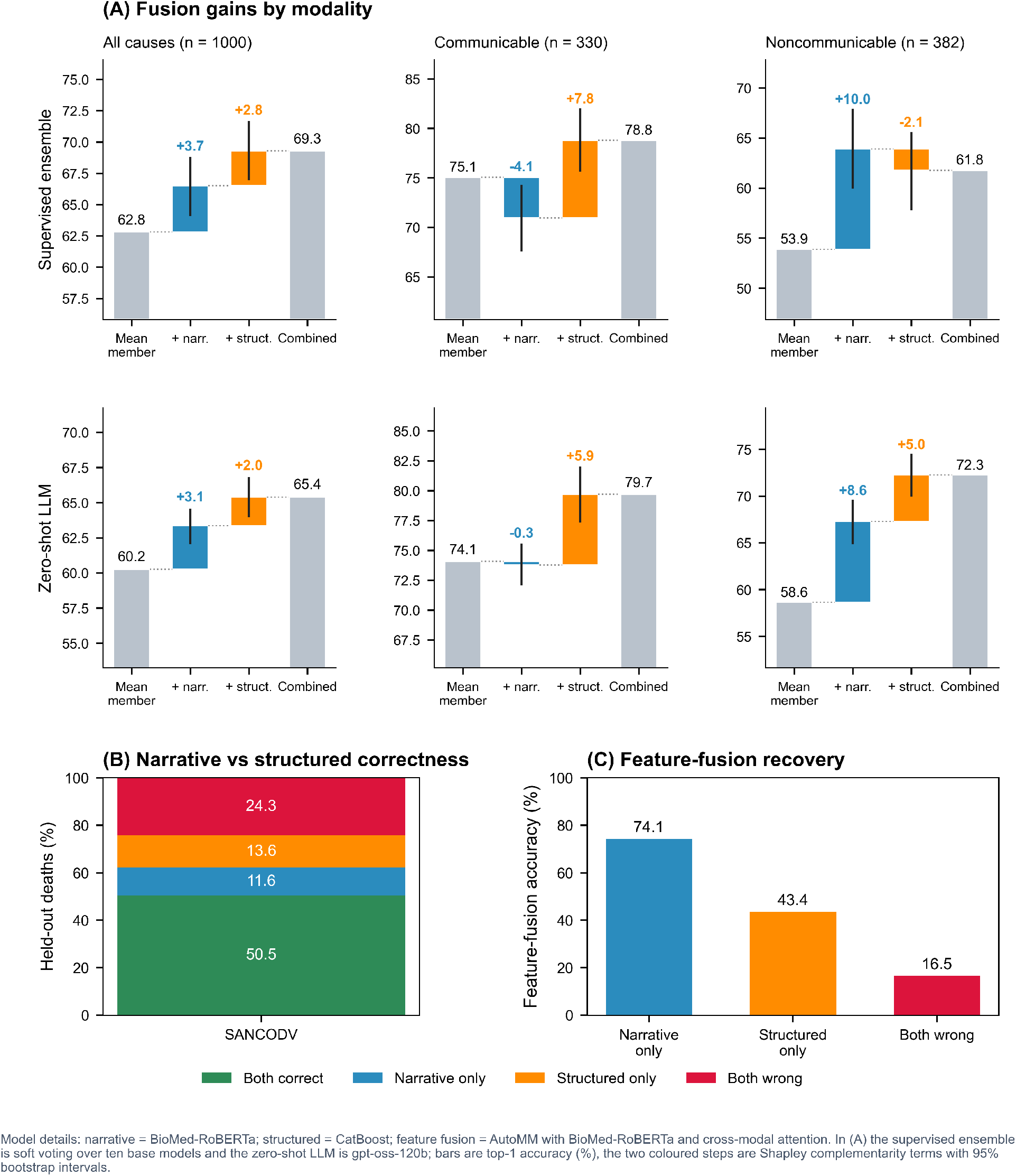
Predictive overlap and fusion gains for narrative and structured inputs. (A) Modality-specific fusion gains by cause group. (B) Overlap in unimodal correctness. (C) Feature-fusion accuracy among deaths misclassified by one or both unimodal models.

BioMed-RoBERTa and CatBoost were both correct for 50.5% of deaths, only BioMed-RoBERTa for 11.6%, only CatBoost for 13.6%, and both were wrong for 24.3% (Figure 2). We measured the cases rescued by the feature-fusion model, which showed the highest balanced accuracy among the fusion pipelines. Feature fusion was correct for 74.1% of the narrative-only cases, 43.4% of the structured-only cases, and 16.5% of the deaths missed by both.

### 3.4 Retrospective calibration

Classwise ECE ranged from 0.010 to 0.025 across the seven representative supervised pipelines, compared with 0.035 for InSilicoVA (Figure 3; Supplementary Table S9). Stacking had the lowest ECE (0.0097), whereas soft voting and class-aware voting shared the lowest class-averaged Brier score (0.0224).

**Figure 3:**
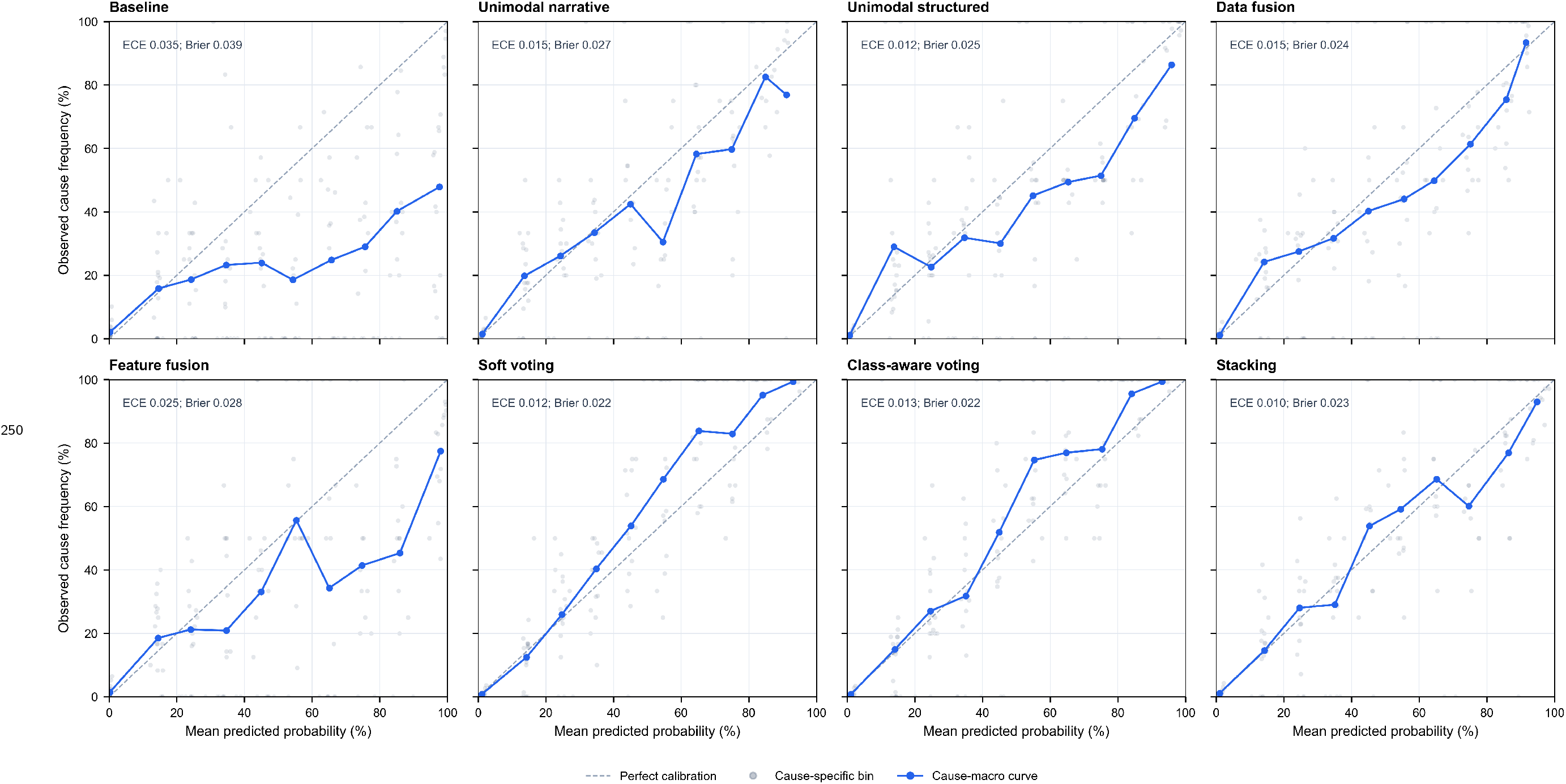
Classwise reliability diagrams. Points are equal-width probability bins for one-vs-rest causes. Lines show class-averaged predicted and observed probabilities. ECE and Brier scores are shown in each panel and summarized in Supplementary Table S9.

### 3.5 Robustness and sensitivity

The performance pattern in Table 2 was consistent across candidate models within each family (Supplementary Section S5). Under coarser and finer cause groupings, the advantage of supervised pipelines over InSilicoVA persisted and the relative ranking of pipelines was similar.

Feature-fusion performance was robust to the choice of narrative backbone but sensitive to the tabular encoder and fusion architecture: in a four-configuration comparison, cross-modal attention raised population-level agreement relative to MLP fusion under either encoder (CSMF accuracy 95.7% vs. 94.2% with categorical embeddings), and only the pairing of categorical embeddings with cross-modal attention separated from the rest on individual-level accuracy.

The nine provinces differ in size and in cause profile, but in the LOPO analysis every supervised pipeline exceeded InSilicoVA in top-1 accuracy in all of them, and in nearly every other comparison. The few exceptions were mostly in CSMF accuracy, chiefly the three decision-fusion pipelines in North West, the province where InSilicoVA’s own CSMF accuracy was highest.

Mean top-1 accuracy was 62.6% for BioMed-RoBERTa, 66.6% for CatBoost, 64.5% for feature fusion, and 68.8–69.2% for the decision-fusion pipelines, versus 51.7% for InSilicoVA, and BioMed-RoBERTa varied more across provinces than CatBoost (top-1 ranges 14.1 vs. 6.5 points). These results support the robustness of models fitted without the target province, with hyperparameters selected globally (Supplementary Section S8.3).

## 4 Discussion

In a national sample of South African VA records, supervised models using narratives or structured responses performed similarly and substantially improved on current best practice— InSilicoVA—for individual COD assignment, with smaller gains in CSMF accuracy. Their relative strengths varied by cause: narratives performed better for noncommunicable diseases and structured responses for communicable diseases, and a similar pattern was observed with the zero-shot LLM. The LLM extended performance on rare causes and achieved the highest balanced accuracy, while the supervised pipelines remained stronger on common causes and population-level agreement. Multimodal fusion further improved performance: among the supervised fusion strategies, decision-level fusion favored overall accuracy, whereas jointly trained fusion yielded larger gains for class-balanced metrics and produced the highest CSMF accuracy point estimate.

Free-text narratives alone contained sufficient diagnostic signal to support cause assignment: narrative-only PLMs performed comparably to models using structured indicators and outperformed InSilicoVA across the individual-level metrics. This is consistent with previous studies reporting narratives containing COD-relevant information.^7,8^ The zero-shot LLM reproduced this result without task-specific fine-tuning or a learned target-population cause distribution, strengthening the evidence that the diagnostic signal lies in the narrative itself rather than solely in supervised fitting. Narratives should therefore be treated as a core input to automated COD estimation rather than reserved for physician review.

The ML models and InSilicoVA used the same harmonized indicators but they rely on different statistical assumptions. InSilicoVA uses a factorized likelihood that treats indicators as conditionally independent given cause, so residual correlation double-counts partially redundant evidence and inflates the resulting posteriors.^5^ Gradient-boosted trees model nonlinear interactions and learn whether an indicator adds information conditional on the others. That difference showed up in both assignments and probabilities: InSilicoVA was the least well calibrated on every calibration measure. The ML advantage in individual assignment did not translate to population estimates, because cause fractions are aggregated from top-1 assignments and misassignments that offset one another leave the distribution unchanged. Changing the estimator alone therefore improves which cause a record receives and how far that assignment can be trusted, without necessarily moving the cause-specific mortality fractions.

Within the supervised models, structured responses were more informative for communicable causes, whereas narratives contributed more for noncommunicable causes. This pattern is consistent with the two modalities describing the same death from different perspectives and at different levels of detail: questionnaires systematically cover prespecified signs and symptoms across respondents, whereas narratives can convey medical history, illness course, care-seeking, and context at a depth generally beyond the scope of a fixed set of questions. The modalities had broadly similar strengths and weaknesses across causes, but differed in which individual deaths they assigned correctly—creating scope for fusion. Several rare causes were consistently challenging and underassigned by supervised pipelines, whereas the zero-shot LLM, which draws on general pretrained knowledge without a data-derived cause-profile prior, recovered more of them even from the same inputs (Supplementary Section S7.4). The LLM’s recovery of more of these causes from each input suggests that both modalities retain diagnostically useful rare-cause information. Sparse labels and a learned study-specific cause distribution may instead draw supervised predictions toward common causes.

Decision fusion produced the largest gain in top-1 accuracy, concentrated among common causes, whereas data and feature fusion gained more on less common causes and therefore on class-balanced metrics (Supplementary Section S7.3). One explanation lies in where the information is combined. Data and feature fusion process both inputs before a cause is assigned, allowing partial evidence from one modality to be weighed against evidence from the other while the decision remains open. Decision fusion instead operates on probability distributions produced after each modality has been processed separately. Among deaths missed by all ten base models, the correct cause often ranked near the top for at least one model but carried little probability relative to a wrong cause consistently supported across models (Supplementary Section S7.4).

Decision fusion therefore consolidated the members’ shared strengths but also preserved their shared errors. Independently trained data- and feature-fusion models recovered some of these deaths by combining the inputs before a cause was assigned, although the broader set of hard cases remained. The similar performance of simple averaging, class-aware weighting, and stacking suggests that simple averaging captured most of the observed decision-fusion gain, consistent with LAVA.^19^ Together, these findings support testing representative fusion strategies against the primary output and operational requirements of the intended deployment.

The zero-shot LLM was competitive with the supervised pipelines for individual COD assignment and, with narratives alone, achieved the highest balanced accuracy, with its advantage concentrated among rare causes. Avoiding a study-derived empirical prior may preserve rare-cause predictions inferred from general medical knowledge while leaving aggregate assignments less closely aligned with population prevalence. Both InSilicoVA and prompted LLMs can in principle operate without labelled deaths, although in this study we adapted InSilicoVA to the customized cause list and trained its symptom–cause probability base rather than using generic values. Prompted LLMs therefore offer an option where labelled deaths are unavailable or rare-cause recognition is important, whereas locally trained supervised pipelines remain preferable when population cause fractions are the primary output.

The error partition identifies data collection as an additional route to improving COD assignment. Feature fusion redistributed errors, recovering some deaths missed by both unimodal pipelines while losing some that one had classified correctly. Shared failures clustered in residual and indeterminate categories (Supplementary Section S7.4). The substantial indeterminate share after physician review further suggests that difficult cases may reflect insufficient diagnostic evidence as well as limitations in model capability. Unlike structured questionnaires, narrative collection lacks comparable protocols and training for probing, elicitation, and recording. A small paired-interviewer study in Malawi found more inter-interviewer discrepancies in open narratives than in closed responses, suggesting that collection practice is one potentially modifiable source of variation.^4^ Future work should assess information sufficiency and model capacity jointly and test whether standardizing narrative collection can help preserve more diagnostic signal for COD assignment.

To our knowledge, this is the first head-to-head comparison of the main unimodal and multimodal VA approaches, including a zero-shot LLM, against the operational algorithm within a single benchmark. All pipelines were evaluated on the same held-out records, reference labels, and metrics, with differences estimated by paired bootstrap. Candidate models and LLM prompt configurations were selected using macro F1 on held-in training data. The feature-fusion architecture and the decision-fusion rules were prespecified. Decision fusion used the same base models, isolating the combination rule from ensemble membership. The inclusion of registered deaths from all nine provinces extends the evidence beyond single-catchment settings. The evaluation covered individual- and population-level performance and calibration, and tested robustness across cause groupings and geographic locations. The open-source implementation supports reproduction and extension.^37^

This study has several limitations. First, physicians assigned reference causes from complete VA records containing both modalities. Fusion gains may reflect both better inference and closer alignment with the evidence used to define the outcome. Physician assignments may also be imperfect, so our metrics measure agreement with this reference rather than with clinical truth. Validation against independently derived causes, such as autopsy or minimally invasive tissue sampling, is needed.^38^ Second, the data are from a high-HIV-prevalence setting in which the remaining causes are relatively rare, preventing separation of data scarcity from model capability. This was especially consequential for learned decision-fusion methods fitted on out-of-fold predictions from smaller training samples. Larger, more balanced datasets are needed to assess their potential. Third, the bootstrap intervals capture held-out sampling variation but not training stochasticity, split selection, or hyperparameter-selection uncertainty. Finally, the sample included deaths at ages 12 years and older from one country. LOPO used globally selected hyperparameters and was therefore not external validation, while the retrospective evaluation could not assess temporal drift, workflow integration, or maintenance costs. External, prospective, and workflow validation across settings, age groups, and languages will support broader implementation.

## 5 Conclusions

Narratives carried substantial diagnostic information that the operational VA algorithms do not use. PLMs using narratives alone matched ML pipelines based on structured responses, while the zero-shot LLM showed particular strength on rare causes without local parameter fitting. These findings support treating narratives as a core input to automated COD systems and using them with the structured questionnaire. Multiple fusion strategies improved individual COD assignment, with gains distributed differently across outcomes, while population-level agreement remained similar because offsetting individual errors muted changes in aggregate cause fractions. These case-level gains could support physician review. Prospective workflow studies should compare representative fusion strategies according to primary outcome of interest and operational capacities, while external validation across settings, age groups, and languages will support broader implementation of multimodal pipelines in routine vital-registration systems.

## Ethics approval and data governance

The South African National Cause of Death Validation (SANCODV) study was approved by the South African Medical Research Council (SAMRC) Human Research Ethics Committee (EC004-2/2017) and reviewed in accordance with U.S. Centers for Disease Control and Prevention human-research protection procedures. Informants provided written consent for participation and use of de-identified data. The present study was a secondary analysis of de-identified records relating to deceased persons, involved no interaction or intervention with living individuals, and did not require additional review by an institutional review board.

## Data availability

The de-identified verbal-autopsy records analyzed in this study are held by SAMRC and are not publicly available, because they contain potentially identifying narrative text about individual deaths. Requests for access should be directed to SAMRC through the corresponding author, and are subject to SAMRC approval, custodian review, and a data use agreement. The derived model outputs needed to reproduce every table and figure in this report, including held-out predictions and bootstrap resample results, are released with the analysis repository.

## Code availability

The core methods, including the zero-shot LLM arm with its prompt, are implemented in the open-source *MultimodalVA* Python package, available at https://github.com/y-chu/MultimodalVA.^37^ Training scripts, model configurations, and trained model checkpoints are released separately at https://github.com/y-chu/MultimodalVA-analysis to support reproduction and downstream use.

## Author contributions

YC conceived the study, designed its methodology, implemented all models, developed the analysis software, curated the analytic data, conducted the analyses, interpreted the results, and drafted the manuscript. SJC contributed to the conceptual framing of the study and supervised the dissertation research. ZW, TM, and RL advised on statistical and methodological aspects. All authors reviewed the manuscript, provided intellectual feedback, and approved the final version.

## Use of AI-assisted technologies

The authors used OpenAI (GPT-4.1 mini), accessed programmatically through the API, solely for conservative copy-editing of narratives—spelling, punctuation and grammar, preserving the original meaning—as part of data preprocessing, as described in the Methods and Supplementary Appendix. The data were provided to the authors de-identified, with direct identifiers removed by the source study (names, addresses and study identifiers), and the authors did not possess a linkage key. The model was not used for classification, inference or generation of study results.

Open-weight generative language models (gpt-oss, MedGemma) were evaluated as the zero-shot arms of the study, as described in the Methods and Supplementary Appendix. They were deployed locally through Ollama to assign causes from the narratives and the structured responses, and their outputs were evaluated alongside the supervised pipelines. They were not used to generate figures or any text in the manuscript.

Claude Code (Anthropic) was used for coding assistance and debugging, for preparing and formatting tables and figures from existing model outputs, and for language editing. The authors reviewed and verified all AI-assisted outputs and take full responsibility for the accuracy and integrity of the work.

## Acknowledgments

We thank Pamela Groenewald and the SANCODV study team for collecting, managing, and sharing the study data. We thank Jason Thomas for technical support with InSilicoVA. We thank the communities in South Africa for their continued support and participation. We thank the Ohio Supercomputer Center for providing computational resources for this research.

