## Supplementary Appendix for "Multimodal Artificial Intelligence for Cause-of-Death Assignment From Verbal Autopsy Records: A National Benchmark Study in South Africa"

### Contents

**S1 Prior narrative and multimodal modeling in verbal autopsy** 3

**S2 Data and preprocessing** 4

**S3 Reference standard and cause categories** 5

**S4 Models and evaluation** 11

**Supplementary references** **59**

This appendix contains additional methodological details, additional results, and interpretation.

### S1 Prior narrative and multimodal modeling in verbal autopsy

Computational research has consistently found cause-of-death (COD) information in verbal-autopsy narratives, using methods ranging from engineered linguistic features to neural, domain-adapted, and contextual representations.[1–6] Most of this work, however, treated the narrative as the primary or sole input rather than examining how it relates to the structured questionnaire.

Studies comparing or combining the two VA components have been fewer, and their findings have varied by dataset, outcome, and fusion design. Early concatenation-based approaches found little aggregate benefit in some settings despite improvements for particular causes,[1,7] whereas models combining learned narrative and questionnaire representations improved on either input alone in another multicause dataset.[8] Decision-level stacking produced more modest gains that varied by age group and metric.[9] These studies show that combining modalities can improve COD assignment, but do not establish a uniform advantage or identify where the gains arise. Cause-specific comparisons have also been limited, making it difficult to determine whether aggregate results conceal systematic differences between the information extracted from narratives and questionnaires.

Recent language-model studies address related but distinct questions. Narrative-based predictions have been used to improve downstream population inference,[10] and GPT models have been evaluated on elicited narratives against algorithms using structured responses,[11] but these comparisons changed both the estimator and its input. Other work converted structured variables into prose rather than using independently elicited narratives, and therefore evaluated a textual representation of questionnaire information rather than multimodal integration.[12] LAVA, published concurrently with the present work, evaluated joint-input language and embedding models, a structured latent-class model, and their ensembles under leave-one-site-out validation.[13] It did not include matched narrative-only and structured-only ablations of the joint-input models or a feature-level architecture with separate modality encoders.

The present study therefore compares narrative-only, structured-only, and data-, feature-, and decision-level fusion on the same records, labels, partitions, and outcomes. This design permits matched evaluation of individual assignment, population-level agreement, calibration, and geographic stability, together with cause-level analyses of where modality and fusion gains occur.

### S2 Data and preprocessing

Verbal autopsies with narratives were de-identified by the source sites before transfer to the investigators, with direct identifiers removed. The investigators did not possess a linkage key.

Thirty-six records without a usable narrative or containing placeholder text only were excluded before the analytic sample was defined. Structured indicators were harmonized to the WHO 2016 VA variables used by the comparator. The de-identified narratives were processed programmatically through the OpenAI API using GPT-4.1 mini solely for conservative copy-editing: spelling, punctuation, grammar, and same-concept shorthand standardization. All digit strings were replaced with positional placeholders locally before transmission and restored afterwards, so that dates, ages, durations, counts, and dosages were never sent to the API. Under the OpenAI API data-use policy in effect at the time of processing, API inputs and outputs were not used to train or improve OpenAI models unless the customer explicitly opted in to such data sharing. The investigators did not opt in.

To minimize alteration of the source information and strictly limit the model's role in preprocessing, the correction prompt permitted only unambiguous changes to spelling, spacing, punctuation, grammar, and same-concept shorthand. It explicitly prohibited diagnostic inference, clinical reinterpretation, changes to negation, numbers, anatomy, or chronology, and expansion of ambiguous abbreviations. The model was not used to infer diagnoses, causes of death, clinical attributes, or other substantive information from the narratives. The full versioned prompt and preprocessing code are provided with the analysis repository. (`https://github.com/y-chu/MultimodalVA-analysis`) Corrected narratives were used for analysis.

Narratives were modest in length (median 95 words, IQR 69–128) (Figure S1) and well within the limits of the models used for analysis. The data-fusion inputs were longer because serialized structured responses were appended.

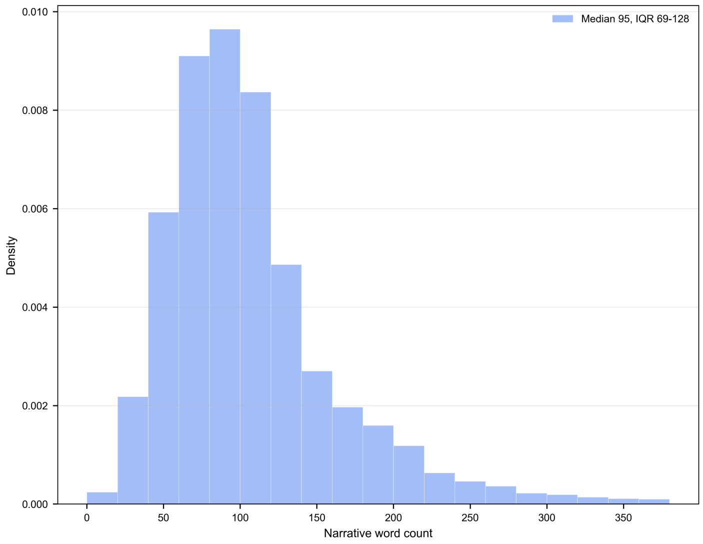

Figure S1: Narrative word-count distribution. Counts are for the corrected narratives used in the primary analysis, after curated shorthand expansion and conservative spelling correction.

#### S3 Reference standard and cause categories

Seventy-five trained physicians reviewed the full WHO 2016 VA record, including the narrative and structured responses. Each record was independently reviewed by two physicians. Disagreements underwent quality-assurance review and adjudication. Physicians reported the underlying

COD and causal sequence on a death certificate, which was coded to ICD-10.[14]

The primary COD grouping contains 19 modeled classes observed among SANCODV deaths aged 12 years or older. Malaria and malnutrition/anaemia had no analytic cases and were not modeled as active classes. Level 1 provides a finer WHO-based grouping where counts permit, and Level 3 collapses causes into six broad groups. The complete mapping from ICD-10 and WHO cause codes to the three analysis levels is given in Table S2, and Table S1 gives the authoritative training and held-out counts for the primary task.

Table S1: Primary cause categories and distributions in the fixed training and held-out partitions. Dashes indicate causes with no SAN-CODV analytic cases.

| Cause of death | Train deaths | Train CSMF (%) | Test deaths | Test CSMF (%) |
|---|---|---|---|---|
| HIV/AIDS | 926 | 23.2 | 232 | 23.2 |
| Pulmonary tuberculosis | 249 | 6.2 | 62 | 6.2 |
| Pneumonia | 43 | 1.1 | 11 | 1.1 |
| Diarrhea | 31 | 0.8 | 7 | 0.7 |
| Meningitis and encephalitis | 19 | 0.5 | 5 | 0.5 |
| Malaria | - | - | - | - |
| Other and unspecified infections | 53 | 1.3 | 13 | 1.3 |
| Neoplasms | 356 | 8.9 | 89 | 8.9 |
| Cardiac disease | 412 | 10.3 | 103 | 10.3 |
| Cerebrovascular disease | 222 | 5.6 | 55 | 5.5 |
| Diabetes | 178 | 4.5 | 45 | 4.5 |
| Chronic pulmonary disease and asthma | 95 | 2.4 | 24 | 2.4 |
| Liver cirrhosis | 50 | 1.3 | 13 | 1.3 |
| Malnutrition and anaemia | - | - | - | - |
| Other and unspecified non communicable diseases | 210 | 5.3 | 53 | 5.3 |
| Maternal conditions | 32 | 0.8 | 8 | 0.8 |
| Traffic / transport accident | 150 | 3.8 | 37 | 3.7 |
| Assault | 185 | 4.6 | 46 | 4.6 |
| Suicide | 58 | 1.5 | 15 | 1.5 |
| Other and unspecified external causes | 104 | 2.6 | 26 | 2.6 |
| Indeterminate | 626 | 15.7 | 156 | 15.6 |

Table S2: Cause-grouping schema. Each analysis category is shown with its ICD-10 codes as recorded in SANCODV and the corresponding WHO cause code, at all three grouping levels.

| Level 3 (broad) | Level 2 (primary) | Level 1 (detailed) | ICD-10 (SANCODV) | WHO cause code |
|---|---|---|---|---|
| Communicable diseases | HIV/AIDS | HIV/AIDS | B20-B24 | 01.03 HIV/AIDS related death |
| | Pulmonary tuberculosis | Pulmonary tuberculosis | A15-A16 | 01.09 Pulmonary tuberculosis |
| | Pneumonia | Pneumonia | J00-J22 | 01.02 Acute resp infect, incl pneumonia |
| | Diarrhea | Diarrhea | A00-A09 | 01.04 Diarrhoeal diseases |
| | Malaria | Malaria | B50-B54 | 01.05 Malaria |
| | Other and unspecified infections | Other and unspecified infections | A40-A41 | 01.01 Sepsis (non-obstetric) |
| | | | B05 | 01.06 Measles |
| | | | A39; G00-G05 | 01.07 Meningitis and encephalitis |
| | | | A33-A35 | 01.08, 10.05 Tetanus |
| | | | A37 | 01.10 Pertussis |
| | | | A90-A99 | 01.11 Haemorrhagic fever |
| | | | A17-A19; A20-A38; A42-A89; B00-B19; B25-B49; B55-B99 | 01.99 Other and unspecified infect dis |
| Non-communicable diseases | Neoplasms | Oral neoplasms | C00-C06 | 02.01 Oral neoplasms |
| | | Digestive neoplasms | C15-C26 | 02.02 Digestive neoplasms |
| | | Respiratory neoplasms | C30-C39 | 02.03 Respiratory neoplasms |
| | | Breast neoplasms | C50 | 02.04 Breast neoplasms |
| | | Reproductive neoplasms M, F | C51-C58; C60-C63 | 02.05, 02.06 Reproductive neoplasms M, F |
| | | Other and unspecified neoplasms | C07-C14; C40-C49; C60-D48 | 02.99 Other and unspecified neoplasms |
| | Diabetes | Diabetes | E10-E14 | 03.03 Diabetes mellitus |
| | Cardiac disease | Acute cardiac disease | I20-I25 | 04.01 Acute cardiac disease |
| | | Other and unspecified cardiac dis | I00-I09; I10-I15; I26-I52; I70-I99 | 04.99 Other and unspecified cardiac dis |
| | Cerebrovascular disease | Stroke | I60-I69 | 04.02 Stroke |

*Continued on next page*

| Level 3 (broad) | Level 2 (primary) | Level 1 (detailed) | ICD-10 (SANCODV) | WHO cause code |
|---|---|---|---|---|
| | Chronic pulmonary disease and asthma | Chronic obstructive pulmonary dis | J40-J44 | 05.01 Chronic obstructive pulmonary dis |
| | | Asthma | J45-J46 | 05.02 Asthma |
| | Liver cirrhosis | Liver cirrhosis | K70-K76 | 06.02 Liver cirrhosis |
| | Other and unspecified non communicable diseases | Other and unspecified NCD | D50-D64 | 03.01 Severe anaemia |
| | | Other and unspecified NCD | D57 | 04.03 Sickle cell with crisis |
| | | Other and unspecified NCD | E40-E46 | 03.02 Severe malnutrition |
| | | Acute abdomen | R10 | 06.01 Acute abdomen |
| | | Renal failure | N17-N19 | 07.01 Renal failure |
| | | Epilepsy | G40-G41 | 08.01 Epilepsy |
| | | Other and unspecified NCD | D55-D89; E00-E07; E15-E35; E50-E90; F00-F99; G06-G09; G10-G37; G50-G99; H00-H95; J30-J39; J47-J99; K00-K31; K35-K38; K40-K93; L00-L99; M00-M99; N00-N16; N20-N99; R00-R09; R11-R94 | 98 Other and unspecified NCD |
| | | Neonatal causes | P05-P07 | 10.01 Prematurity |
| | | | P20-P22 | 10.02 Birth asphyxia |
| | | | P23-P25 | 10.03 Neonatal pneumonia |
| | | | P36 | 10.04 Neonatal sepsis |
| | | | Q00-Q99 | 10.06 Congenital malformation |
| | | | P00-P04; P08-P15; P26-P35; P37-P94; P96 | 10.99 Other and unspecified neonatal CoD |
| | | | P95 | 11.01 Fresh stillbirth |
| | | | P95 | 11.02 Macerated stillbirth |
| Injuries | Traffic / transport accident | Traffic / transport accident | V01-V89 | 12.01 Road traffic accident |
| | | | V90-V99 | 12.02 Other transport accident |
| | Assault | Assault | X85-Y09 | 12.09 Assault |
| | Suicide | Suicide | X60-X84 | 12.08 Intentional self-harm |
| | Other and unspecified external causes | Accid fall | W00-W19 | 12.03 Accid fall |

*Continued on next page*

| Level 3 (broad) | Level 2 (primary) | Level 1 (detailed) | ICD-10 (SANCODV) | WHO cause code |
|---|---|---|---|---|
| | | Accid drowning and submersion | W65-W74 | 12.04 Accid drowning and submersion |
| | | Other and unspecified external CoD | X00-X19 | 12.05 Accid expos to smoke, fire & flame |
| | | | X20-X29 | 12.06 Contact with venomous plant/animal |
| | | | X40-X49 | 12.07 Accid poisoning and noxious subs |
| | | | X30-X39 | 12.10 Exposure to force of nature |
| | | | S00-T99; W20-W64; W75-W99; X50-X59; Y10-Y98 | 12.99 Other and unspecified external CoD |
| Maternal conditions | Maternal conditions | Maternal conditions | O00 | 09.01 Ectopic pregnancy |
| | | | O03-O08 | 09.02 Abortion-related death |
| | | | O10-O16 | 09.03 Pregnancy-induced hypertension |
| | | | O46; O67; O72 | 09.04 Obstetric haemorrhage |
| | | | O63-O66 | 09.05 Obstructed labour |
| | | | O85; O75.3 | 09.06 Pregnancy-related sepsis |
| | | | O99.0 | 09.07 Anaemia of pregnancy |
| | | | O71 | 09.08 Ruptured uterus |
| | | | O01-O02; O20-O45; O47-O62; O68-O70; O73-O84; O86-O99 | 09.99 Other and unspecified maternal CoD |
| Indeterminate | Indeterminate | Indeterminate | R95-R99 | 99 Cause of death unknown |

### S4 Models and evaluation

Figure S2 summarizes the model inputs and the points at which the two modalities are combined. The core methods are implemented in the open-source *MultimodalVA* Python package.[15]

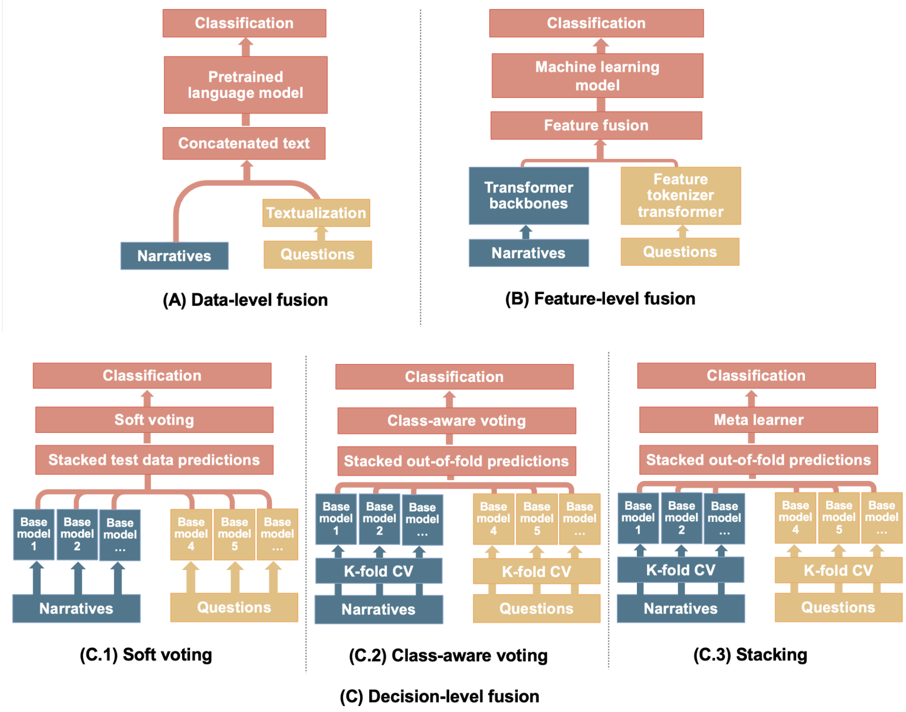

Figure S2: Multimodal fusion strategies. Data fusion concatenates serialized structured responses with the narrative; feature fusion combines learned modality representations; decision fusion combines predicted probability vectors.

#### S4.1 Candidate models and checkpoints

Narrative candidates were BERT, BlueBERT, BioClinicalBERT, ClinicalBERT, BiomedBERT, and BioMed-RoBERTa. Structured candidates were LightGBM, CatBoost, XGBoost, gradient-boosted decision trees, random forest, and a multilayer perceptron. Data-fusion candidates were ClinicalBigBird and ClinicalLongformer. Feature fusion was implemented in AutoGluon AutoMM. The two retained feature-fusion variants both used BioMed-RoBERTa, with and without author-specified cross-modal attention. Table S3 records public PLM checkpoints.

Table S3: Pretrained language models used in narrative and data-fusion pipelines.

| **Model** | **Checkpoint** |
|---|---|
| ClinicalBERT | `medicalai/ClinicalBERT` |
| BioClinicalBERT | `emilyalsentzer/Bio_ClinicalBERT` |
| BlueBERT | `bionlp/bluebert_pubmed_mimic_uncased_L-12_H-768_A-12` |
| BiomedBERT | `microsoft/BiomedNLP-BiomedBERT-base-uncased-abstrac t-fulltext` |
| BioMed-RoBERTa | `allenai/biomed_roberta_base` |
| BERT | `bert-base-uncased` |
| ClinicalBigBird | `yikuan8/Clinical-BigBird` |
| ClinicalLongformer | `yikuan8/Clinical-Longformer` |

#### S4.2 Fusion definitions

Data fusion serialized the structured questionnaire into natural-language sentences and appended them to the narrative, so that one model read a single fused text. The narrative always comes first and the serialized questionnaire second, so that any truncation at the maximum sequence length falls on the structured sentences rather than on the narrative. Demographic items were rendered as one opening sentence and each remaining item as a short affirmative or negative clause. The fused record exceeds the 512-token limit of standard PLMs, so the candidates were long-context checkpoints, ClinicalLongformer and ClinicalBigBird, with a maximum sequence length of 1,346 tokens set at the 95th percentile of fused-text length. This strategy requires no architectural change: the model sees both modalities as one document, but the structured responses lose their tabular form.

Feature fusion kept the two modalities in their native form and combined them inside one model, implemented in AutoGluon AutoMM. Three choices define a configuration.

- *Narrative backbone.* We used the narrative model with the highest training-CV macro F1,

BioMed-RoBERTa, so that the fused model and the narrative-only pipeline share an encoder.

- *Structured encoder.* We evaluated two: a categorical and numerical embedding of each questionnaire item, and a feature-tokenizer transformer that attends across items.
- *Fusion module.* When cross-modal attention was specified, a fusion transformer applied attention over the joint sequence of narrative and structured representations. When it was not, the default multilayer perceptron combined the two representations by concatenation.

All configurations used the `best_quality` preset with default model settings and the same 24-hour training budget, without per-configuration hyperparameter search, so they form a controlled architecture contrast.

For model $m$ and case $i$, let $\mathbf{p}_i^{(m)}$ be the probability vector over $C = 19$ causes. Soft voting used

$$\mathbf{p}_i^{\text{soft}} = M^{-1} \sum_{m=1}^{M} \mathbf{p}_i^{(m)}.$$

Its pool contained five narrative PLMs (BERT, BlueBERT, BioClinicalBERT, BiomedBERT, and BioMed-RoBERTa) and five structured models (LightGBM, CatBoost, XGBoost, GBDT, and a multilayer perceptron). InSilicoVA was excluded: it is the comparator against which the ensembles are evaluated.

All three decision-fusion pipelines drew on this same set of ten models, so that any difference between them reflects the combination rule rather than the members. They differ in which fit of those models they use: soft voting averaged the probabilities of each unimodal pipeline's own fitted model, whereas class-aware voting and stacking learned their combination rules from out-of-fold predictions obtained by refitting the ten models within the training partition, where each fold is fitted on four-fifths of that partition.

Class-aware voting used five-fold out-of-fold (OOF) predictions to estimate a model-by-cause reliability matrix from one-vs-rest F1 scores. We used per-cause F1 because it gives equal weight to a model's ability to identify a cause and to its tendency to over-assign it, which is the balance required when most causes are of low prevalence and models must still be distinguishable from one another. We chose it on these grounds rather than on observed held-out performance. Class-specific weights were shrunk toward $1/M$ with a base shrinkage of 0.1, increasing toward $1/M$ as cause support decreased, then gated for each case by the class on which each model placed its largest probability. The normalized gates formed a convex combination of the base probabilities. We compared the learned weights with equal weighting by three-fold cross-validation on the same OOF matrix and retained equal weights where the learned weights did not improve macro F1, so that the pipeline reduces to soft voting on the same base models wherever cause-varying reliability is not supported by the training data.

Stacking used the same ten base models. The OOF probability vectors were concatenated as meta-features. Multinomial logistic regression, LightGBM, XGBoost, and random forest meta-learners with fixed candidate configurations were compared by stratified cross-validated macro F1 on the OOF matrix. The chosen meta-learner was refit on the full OOF matrix. The held-out test set was used only once for final evaluation.[16]

#### S4.3 Model configuration and hyperparameter optimization

Optuna searches using the Tree-structured Parzen Estimator sampler were run separately for each candidate narrative, structured, and data-fusion model, using the training partition only.[17,18] The objective was mean macro F1 in stratified 3-fold cross-validation. Table S4 summarizes the trial budgets and pruning rules. The selected configuration for each candidate was refit on the complete training partition.

Within the narrative, structured, and data-fusion families, the representative reported in Table 2 was the candidate with the highest training-partition cross-validated macro F1: BioMed-RoBERTa, CatBoost, and ClinicalLongformer, respectively. Feature fusion used BioMed-RoBERTa on the same criterion, but its structured encoder and fusion module were prespecified rather than selected by Optuna. The decision-fusion methods reused the selected configurations of their base models. Their combination rules were also prespecified.

Table S4: Hyperparameter-search design.

| **Family** | **Trials run** | **Trials done** | **Validation and objective** | **Search control** |
|---|---|---|---|---|
| Narrative PLMs | 50 | 27–38 | Stratified 3-fold CV<br>Macro F1 | TPE sampler<br>Median pruner<br>Patience 4 |
| Structured ML | 60 | 60 | Stratified 3-fold CV<br>Macro F1 | TPE sampler<br>No pruner |
| Data-fusion PLMs | 40 | 27–29 | Stratified 3-fold CV<br>Macro F1 | TPE sampler<br>Median pruner<br>Patience 4 |
| Feature fusion | – | – | Prespecified architecture<br>No search | AutoMM `best_quality` preset<br>24-hour budget |

The search spaces and selected values for all tunable candidates are reported in Table S5.

Table S5: Hyperparameter search spaces and selected values.

| Parameter | Search space | Selected value |
|---|---|---|
| **BERT** | | |
| Learning rate | $5.000 \times 10^{-6}$ to $4.000 \times 10^{-5}$ (log scale) | $3.974 \times 10^{-5}$ |
| Batch size | 8, 16, 32 | 8 |
| Epochs | 3, 5 | 5 |
| Weight decay | 0.01 to 0.3 | 0.106 |
| Warm-up ratio | 0 to 0.2 | 0.118 |
| Gradient accumulation | 1, 2 | 2 |
| Frozen encoder layers | 2, 4, 6 | 4 |
| Classifier dropout | 0.2 to 0.5 | 0.326 |
| Label smoothing | 0.05 to 0.15 | 0.123 |
| **BioClinicalBERT** | | |
| Learning rate | $5.000 \times 10^{-6}$ to $4.000 \times 10^{-5}$ (log scale) | $3.940 \times 10^{-5}$ |
| Batch size | 8, 16, 32 | 8 |
| Epochs | 3, 5 | 5 |
| Weight decay | 0.01 to 0.3 | 0.073 |
| Warm-up ratio | 0 to 0.2 | 0.178 |
| Gradient accumulation | 1, 2 | 1 |
| Frozen encoder layers | 2, 4, 6 | 2 |
| Classifier dropout | 0.2 to 0.5 | 0.345 |
| Label smoothing | 0.05 to 0.15 | 0.09 |
| **BiomedBERT** | | |

*Continued on next page*

| Parameter | Search space | Selected value |
| --- | --- | --- |
| Learning rate | $5.000 \times 10^{-6}$ to $4.000 \times 10^{-5}$ (log scale) | $3.430 \times 10^{-5}$ |
| Batch size | 8, 16, 32 | 8 |
| Epochs | 3, 5 | 5 |
| Weight decay | 0.01 to 0.3 | 0.022 |
| Warm-up ratio | 0 to 0.2 | 0.172 |
| Gradient accumulation | 1, 2 | 1 |
| Frozen encoder layers | 2, 4, 6 | 2 |
| Classifier dropout | 0.2 to 0.5 | 0.228 |
| Label smoothing | 0.05 to 0.15 | 0.051 |
| **BlueBERT** | | |
| Learning rate | $5.000 \times 10^{-6}$ to $4.000 \times 10^{-5}$ (log scale) | $3.462 \times 10^{-5}$ |
| Batch size | 8, 16, 32 | 8 |
| Epochs | 3, 5 | 5 |
| Weight decay | 0.01 to 0.3 | 0.033 |
| Warm-up ratio | 0 to 0.2 | 0.173 |
| Gradient accumulation | 1, 2 | 1 |
| Frozen encoder layers | 2, 4, 6 | 2 |
| Classifier dropout | 0.2 to 0.5 | 0.317 |
| Label smoothing | 0.05 to 0.15 | 0.129 |
| **ClinicalBERT** | | |
| Learning rate | $5.000 \times 10^{-6}$ to $4.000 \times 10^{-5}$ (log scale) | $3.095 \times 10^{-5}$ |
| Batch size | 8, 16, 32 | 8 |
| Epochs | 3, 5 | 5 |
| Weight decay | 0.01 to 0.3 | 0.011 |
| Warm-up ratio | 0 to 0.2 | 0.058 |

*Continued on next page*

| Parameter | Search space | Selected value |
| --- | --- | --- |
| Gradient accumulation | 1, 2 | 1 |
| Frozen encoder layers | 2, 4, 6 | 6 |
| Classifier dropout | 0.2 to 0.5 | – |
| Label smoothing | 0.05 to 0.15 | 0.136 |
| **BioMed-RoBERTa** | | |
| Learning rate | $5.000 \times 10^{-6}$ to $4.000 \times 10^{-5}$ (log scale) | $3.179 \times 10^{-5}$ |
| Batch size | 8, 16, 32 | 8 |
| Epochs | 3, 5 | 5 |
| Weight decay | 0.01 to 0.3 | 0.241 |
| Warm-up ratio | 0 to 0.2 | 0.181 |
| Gradient accumulation | 1, 2 | 2 |
| Frozen encoder layers | 2, 4, 6 | 4 |
| Classifier dropout | 0.2 to 0.5 | 0.286 |
| Label smoothing | 0.05 to 0.15 | 0.139 |
| **ClinicalBigBird** | | |
| Learning rate | $5.000 \times 10^{-6}$ to $4.000 \times 10^{-5}$ (log scale) | $2.982 \times 10^{-5}$ |
| Batch size | 1, 2, 4 | 1 |
| Epochs | 3, 5 | 5 |
| Weight decay | 0.01 to 0.3 | 0.219 |
| Warm-up ratio | 0 to 0.2 | 0.116 |
| Gradient accumulation | 4, 8, 16 | 4 |
| Frozen encoder layers | 2, 4 | 4 |
| Classifier dropout | 0.2 to 0.5 | 0.357 |
| Label smoothing | 0.05 to 0.15 | 0.125 |
| **ClinicalLongformer** | | |

*Continued on next page*

| Parameter | Search space | Selected value |
|---|---|---|
| Learning rate | $5.000 \times 10^{-6}$ to $4.000 \times 10^{-5}$ (log scale) | $3.934 \times 10^{-5}$ |
| Batch size | 1, 2, 4 | 2 |
| Epochs | 3, 5 | 5 |
| Weight decay | 0.01 to 0.3 | 0.205 |
| Warm-up ratio | 0 to 0.2 | 0.198 |
| Gradient accumulation | 4, 8, 16 | 4 |
| Frozen encoder layers | 2, 4 | 4 |
| Classifier dropout | 0.2 to 0.5 | – |
| Label smoothing | 0.05 to 0.15 | 0.111 |
| **CatBoost** | | |
| Iterations | 150 to 1,440 | 1,220 |
| Learning rate | 0.005 to 0.15 (log scale) | 0.149 |
| Tree depth | 4 to 9 | 9 |
| $L_2$ leaf regularization | 0.01 to 30 (log scale) | 14.569 |
| Boosting type | Ordered, Plain | Ordered |
| **GBDT** | | |
| Estimators | 100 to 840 | 710 |
| Learning rate | 0.005 to 0.15 (log scale) | 0.018 |
| Max depth | 2 to 6 | 2 |
| Min samples split | 2 to 50 | 50 |
| Min samples leaf | 1 to 20 | 16 |
| Subsample | 0.5 to 1 | 0.867 |
| **LightGBM** | | |
| Estimators | 150 to 1,440 | 1,389 |
| Learning rate | 0.005 to 0.15 (log scale) | 0.079 |

*Continued on next page*

| Parameter | Search space | Selected value |
|---|---|---|
| Max depth | -1, 4, 6, 8, 10 | 10 |
| Leaves | 31 to 152 | 73 |
| Min child samples | 10 to 150 | 40 |
| Subsample | 0.6 to 1 | 0.944 |
| Column subsample | 0.3 to 0.9 | 0.34 |
| $L_1$ regularization | $1.000 \times 10^{-4}$ to 20 (log scale) | 1.056 |
| $L_2$ regularization | $1.000 \times 10^{-4}$ to 30 (log scale) | 0.524 |
| **MLP** | | |
| Hidden layer sizes | (128,), (256,), (256, 128), (512, 256), (256, 128, 64) | [256, 128] |
| Activation | tanh, relu | tanh |
| Initial learning rate | $1.000 \times 10^{-4}$ to 0.05 (log scale) | $1.304 \times 10^{-4}$ |
| $L_2$ penalty | $1.000 \times 10^{-6}$ to 0.1 (log scale) | 0.026 |
| Batch size | 32, 64, 128, 256 | 128 |
| **Random forest** | | |
| Estimators | 150 to 960 | 377 |
| Max depth | None, 10, 20, 30 | None |
| Min samples split | 2 to 50 | 10 |
| Min samples leaf | 1 to 20 | 1 |
| Max features | sqrt, log2, 0.1, 0.2, 0.5 | 0.5 |
| **XGBoost** | | |
| Estimators | 150 to 1,440 | 1,115 |
| Learning rate | 0.005 to 0.15 (log scale) | 0.016 |
| Max depth | 3 to 8 | 7 |
| Subsample | 0.6 to 1 | 0.726 |
| Column subsample | 0.3 to 0.9 | 0.769 |

*Continued on next page*

| Parameter | Search space | Selected value |
| --- | --- | --- |
| Gamma | 0 to 5 | 0.628 |
| Min child weight | 1 to 8 | 3 |
| $L_1$ regularization | $1.000 \times 10^{-4}$ to 20 (log scale) | $8.023 \times 10^{-4}$ |
| $L_2$ regularization | $1.000 \times 10^{-4}$ to 30 (log scale) | 3.741 |

##### S4.3.1 InSilicoVA configuration and sensitivity analysis

InSilicoVA was fitted through `insilico.train`. Symptom-given-cause probabilities were estimated from the same training partition used by the supervised models rather than taken from the generic probability base. At a missingness threshold of 0.95, 131 symptoms were removed. Seventy symptom–cause combinations absent from the training data were imputed using mean symptom prevalence. The same preprocessing was used for every specification.

The reported specification assigned the estimated probabilities to ranked levels using fixed cut-points and re-estimated the level values during sampling, the specification its developers recommend.[19] It was fixed in advance and used at every cause-grouping level.

Two sensitivity analyses varied it (Table S6). The first compared the reported model with one that holds the estimated probabilities fixed and one that uses quantile-based rather than fixed cut-points. The second held the specification fixed and varied the chain length. Exact settings are in the table footnote.

Specification had a larger effect than chain length once convergence was reached. Holding the estimated probabilities fixed improved held-out top-1 and CSMF accuracy, whereas quantile-based levels performed less well. All supervised candidates nevertheless remained more accurate than the no-updating specification on each of the five individual-level metrics. Fourteen of 19 had lower CSMF accuracy, by at most 4.5 percentage points. Runs with 10,000 to 40,000 simulations passed the package convergence check and produced similar results. The 4,000-simulation run did not converge and was less accurate.

Table S6: InSilicoVA sensitivity to probability-level specification and MCMC settings. Panel A compares specifications. Cross-validated macro F1 is from 3-fold training-partition runs, and the remaining metrics are from held-out evaluation using the primary MCMC settings. Panel B varies chain length under the reported specification without automatic extension. The asterisk marks the specification used throughout. *Settings.* Reported specification: `type = "fixed"`, `updateCondProb = TRUE`, `keepProbbase.level = TRUE`, `levels.strength = 1`; primary run 20,000 simulations, 10,000 burn-in, thinning 20, automatic chain extension, seed 42. Panel A rows 2 and 3: probabilities held fixed, `type = "empirical"`, `updateCondProb = FALSE`; quantile levels, `type = "quantile"`, `updateCondProb = TRUE`, `levels.strength = 0.01`. All rows keep `keepProbbase.level = TRUE`.

| | CV macro F1 (%, SD) | Top-1 (%) | Balanced accuracy (%) | Macro F1 (%) | CSMF (%) | CCCSMF (%) |
|---|---|---|---|---|---|---|
| *A. Specification* | | | | | | |
| Fixed levels, updated* | 41.7 (1.5) | 53.6 | 43.1 | 41.1 | 88.5 | 68.9 |
| No updating | 41.7 (1.9) | 55.7 | 43.0 | 41.9 | 93.1 | 81.2 |
| Quantile levels, updated | 36.1 (0.2) | 48.4 | 39.9 | 34.2 | 81.4 | 49.5 |
| *B. Chain length, reported specification* | | | | | | |
| Nsim 4,000, burn-in 2,000, thin 10 | – | 52.0 | 43.1 | 40.2 | 86.8 | 64.2 |
| Nsim 10,000, burn-in 5,000, thin 20 | – | 53.6 | 43.1 | 41.2 | 88.6 | 69.1 |
| Nsim 20,000, burn-in 10,000, thin 20 | – | 53.6 | 43.1 | 41.1 | 88.5 | 68.9 |
| Nsim 40,000, burn-in 20,000, thin 40 | – | 53.6 | 42.9 | 41.2 | 88.8 | 69.7 |

#### S4.4 Generative large language model

An open-weight generative language model assigned one underlying cause from the same 19-cause list without parameter updates. Four candidate models were evaluated on a 300-record development subset of the training partition: `gpt-oss:120b`, `gpt-oss:20b`, `medgemma:27b`, and `medgemma1.5:latest`. The model with the highest macro F1, `gpt-oss:120b`, was retained. Models were served locally through Ollama with temperature 0, seed 42, and low reasoning effort. The narrative, structured, and combined arms received, respectively, the copy-edited narrative, the structured responses rendered as text, or both. Each arm subsequently scored all 1,000 held-out records.

The prompt comprised a common task instruction and four optional modules. M1 provided South African study context while cautioning against inferring individual causes from population prevalence; M2 provided a reasoning scaffold distinguishing the illness sequence, mode of dying, immediate cause, underlying cause, and contributing conditions; M3 defined the scope of the 19 cause labels using WHO VA categories and ICD-10 ranges; and M4 provided cross-cause arbitration rules condensed from WHO physician-review guidance. Module combinations were selected separately for each input by macro F1 on the development subset (Table S7). The complete versioned prompts and configuration files are available in the code release (`https://github.com/y-chu/MultimodalVA-analysis`).

Table S7: Prompt configurations evaluated on the 300-record development subset.

| Modules | Input | Macro F1 | Top-1 accuracy | CSMF accuracy | Prompt tokens |
|---|---|---|---|---|---|
| M1, M3* | Narrative | 0.534 | 61.0 | 87.9 | 2,552 |
| None | Narrative | 0.533 | 59.3 | 84.9 | 1,398 |
| M1, M2, M3, M4 | Narrative | 0.495 | 60.7 | 85.6 | 3,868 |
| M1, M2, M3 | Narrative | 0.471 | 59.0 | 84.9 | 2,646 |
| M2, M3, M4 | Narrative | 0.500 | 60.3 | 82.2 | 3,803 |
| M1, M2, M3, M4* | Narrative + structured | 0.506 | 62.7 | 82.2 | 5,084 |
| M1, M3, M4 | Narrative + structured | 0.506 | 62.3 | 81.2 | 5,015 |
| M1, M3 | Narrative + structured | 0.464 | 58.0 | 76.5 | 3,766 |
| M1, M3, M4* | Structured only | 0.489 | 59.7 | 82.6 | 4,909 |
| M1, M3 | Structured only | 0.425 | 51.3 | 75.2 | 3,647 |

*Note.* Top-1 and CSMF accuracies are percentages. An asterisk marks the configuration selected for each input. All runs used `gpt-oss:120b` with identical decoding settings, and none produced a schema-invalid response. An alternative wording of M1 and two few-shot prompts were also evaluated but not selected. The alternative wording achieved a macro F1 of 0.527, and the best few-shot configuration achieved 0.533, compared with 0.534 for the selected narrative configuration. The two few-shot prompts contained 6,865 and 8,026 tokens, compared with 2,552 for the selected configuration.

#### S4.5 Evaluation metrics

We evaluated performance at both the individual and the population level. Let $C$ be the number of causes, $N$ the number of held-out deaths, $y_i$ the reference cause for death $i$, and $\hat{y}_i$ the cause the model assigned. True positives (TP), false positives (FP), true negatives (TN), and false negatives (FN) are read from the confusion matrix for one cause treated as a one-versus-rest outcome.

##### Individual-level evaluation

- **Top-1 accuracy:** the proportion of deaths whose reference cause was the cause the model ranked first.
- **Top-3 accuracy:** the proportion of deaths whose reference cause was among the three causes with the largest predicted probabilities.
- **Precision, recall, and F1:** within a cause, $TP/(TP+FP)$, $TP/(TP+FN)$, and their harmonic mean.
- **Balanced accuracy:** the unweighted mean of cause-specific recall, which gives every cause equal weight regardless of how many deaths it contains.

$$\text{Balanced accuracy} = \frac{1}{C}\sum_{c=1}^{C} \text{Recall}_c$$

Per-cause measures were aggregated in two ways:

- *Macro:* the unweighted mean across causes, so a rare cause counts as much as a common one.

$$\text{Macro F1} = \frac{1}{C}\sum_{c=1}^{C} \text{F1}_c$$

- *Weighted:* the mean across causes weighted by the number of held-out deaths $n_c$ in each cause.

$$\text{Weighted F1} = \sum_{c=1}^{C} \frac{n_c}{N}\text{F1}_c$$

**Population-level evaluation**

- **Cause-specific mortality fraction (CSMF) accuracy:** how closely the predicted cause distribution matches the reference distribution, where $t_c$ and $q_c$ are the reference and predicted fractions for cause $c$, the latter computed from the pipeline's top-1 assignments.[20] It measures net agreement across the population, so misclassifications that offset one another do not penalize the score.

$$\text{CSMF accuracy} = 1 - \frac{\sum_{c=1}^{C} |q_c - t_c|}{2\,(1 - \min_c t_c)}$$

We took $q_c$ from each pipeline's top-1 assignments rather than by averaging its predicted probabilities across deaths. A model that returned the training cause frequencies for every death, and so carried no information about which death had which cause, would reproduce the reference fractions almost exactly. Under probability aggregation, CSMF accuracy rewards agreement with the marginal distribution rather than correct assignment, and systematically favors models whose probabilities are shrunk toward that distribution over those that commit to a cause. Taking cause fractions from top-1 assignments avoids this, because a model that returned the same distribution for every death would then place every death in one cause and reproduce the reference fractions poorly.

- **Chance-corrected CSMF (CCCSMF) accuracy:** CSMF accuracy adjusted for the agreement expected from random allocation. We used 0.632 as the mean of random allocation, following the convention of Flaxman et al.[21] Because that constant is fixed while the empirical chance level depends on the number of causes, we report CCCSMF at the primary 19-cause level only.

$$\text{CCCSMF accuracy} = \frac{\text{CSMF accuracy} - 0.632}{1 - 0.632}$$

##### Calibration

Calibration asks whether a predicted probability matches the frequency with which the cause occurs. We treated each cause as a one-versus-rest outcome, placed the predicted probabilities $p_{ic}$ into $B = 10$ equal-width bins, and compared the mean predicted probability $\bar{p}_{cb}$ with the observed frequency $\bar{o}_{cb}$ in each bin $b$ of size $n_{cb}$ for cause $c$.

- **Classwise expected calibration error (ECE):** the bin-size-weighted mean absolute gap between predicted and observed frequency within a cause, averaged equally across causes. Lower is better.

$$\mathrm{ECE} = \frac{1}{C}\sum_{c=1}^{C}\sum_{b=1}^{B}\frac{n_{cb}}{N}\left|\bar{o}_{cb} - \bar{p}_{cb}\right|$$

- **Maximum calibration error (MCE):** the largest such gap over all nonempty cause-bins. Because it is determined by a single bin, it is unstable where causes are sparse and we do not use it to rank pipelines.

$$\mathrm{MCE} = \max_{c,b}\left|\bar{o}_{cb} - \bar{p}_{cb}\right|$$

- **Brier score:** the mean squared difference between the predicted probability and the outcome, averaged over all deaths and causes. It rewards probabilities that are both well calibrated and confident. Lower is better.

$$\text{Brier score} = \frac{1}{NC}\sum_{i=1}^{N}\sum_{c=1}^{C}\left(p_{ic} - \mathbb{1}[y_i = c]\right)^2$$

### S5 Complete held-out results

The full leaderboard includes all narrative, structured, data-fusion candidates, BioMed-RoBERTa AutoMM feature-fusion variants, all decision-level methods, and the three large-language-model arms, marked † and placed in the block whose input column each one reads. It presents point estimates along with 95% percentile-bootstrap confidence intervals (1,000 resamples) for all metrics.

Table S8: Full model performance leaderboard, held-out test set ($n = 1{,}000$), as point estimates with 95% bootstrap confidence intervals. Bold marks the highest value within a metric, descriptively. Rows marked † are the LLM arms of Section S4.4.

| Model | Top-1 accuracy | Top-3 accuracy | Balanced accuracy | Weighted F1 | Macro F1 | CSMF accuracy | CCCSMF accuracy |
|---|---|---|---|---|---|---|---|
| Baseline (InSilicoVA) | 53.6 (50.4 to 56.5) | 78.5 (75.9 to 80.8) | 43.1 (39.1 to 47.1) | 53.8 (50.5 to 56.9) | 41.1 (37.3 to 44.1) | 88.5 (84.8 to 90.1) | 68.9 (58.6 to 73.0) |
| **Unimodal narrative** | | | | | | | |
| BlueBERT | 61.5 (58.4 to 64.2) | 83.5 (81.4 to 85.8) | 49.2 (45.6 to 52.4) | 60.3 (57.1 to 63.2) | 47.8 (44.3 to 50.4) | 93.8 (90.0 to 94.8) | 83.1 (72.8 to 85.8) |
| BioClinicalBERT | 63.0 (59.9 to 65.9) | 83.4 (81.0 to 85.7) | 49.6 (45.9 to 52.8) | 61.7 (58.6 to 64.8) | 48.8 (45.2 to 51.7) | 94.1 (90.7 to 94.8) | 83.9 (74.7 to 85.8) |
| BioMed-RoBERTa | 62.1 (59.0 to 64.9) | 84.8 (82.6 to 86.9) | 49.2 (45.6 to 52.4) | 60.7 (57.7 to 63.7) | 47.8 (44.3 to 50.5) | 92.3 (89.0 to 93.6) | 79.0 (70.2 to 82.6) |
| BiomedBERT | 63.7 (60.5 to 66.5) | 83.2 (80.8 to 85.4) | 52.2 (48.0 to 55.8) | 62.4 (59.3 to 65.4) | 51.3 (47.0 to 54.1) | 92.7 (89.1 to 94.0) | 80.1 (70.3 to 83.6) |
| ClinicalBERT | 62.3 (59.3 to 65.2) | 82.7 (80.5 to 85.0) | 49.3 (45.8 to 52.6) | 60.9 (57.7 to 63.8) | 48.8 (45.2 to 51.5) | 92.4 (88.8 to 93.4) | 79.2 (69.5 to 82.0) |
| BERT | 63.5 (60.6 to 66.4) | 82.8 (80.5 to 85.0) | 50.6 (47.2 to 53.9) | 62.0 (59.0 to 65.0) | 49.3 (45.6 to 51.9) | 93.3 (89.7 to 94.4) | 81.7 (71.9 to 84.7) |
| gpt-oss-120b† | 61.3 (58.2 to 64.2) | 77.3 (74.6 to 80.0) | **60.6 (55.2 to 65.7)** | 61.9 (58.8 to 64.9) | 56.3 (51.2 to 59.9) | 85.6 (82.5 to 87.9) | 60.9 (52.4 to 67.0) |
| **Unimodal structured** | | | | | | | |
| LightGBM | 63.7 (60.8 to 66.6) | 87.0 (85.1 to 89.1) | 49.4 (45.6 to 53.2) | 62.6 (59.5 to 65.7) | 50.6 (46.8 to 53.6) | 92.8 (89.5 to 94.1) | 80.3 (71.6 to 83.9) |
| CatBoost | 64.1 (61.2 to 66.9) | 87.8 (85.8 to 89.8) | 50.2 (46.1 to 54.6) | 62.9 (59.8 to 65.9) | 52.0 (47.1 to 55.5) | 92.1 (88.6 to 93.3) | 78.4 (69.1 to 81.7) |
| XGBoost | 64.6 (61.6 to 67.6) | 87.3 (85.3 to 89.2) | 49.9 (46.5 to 53.5) | 63.2 (60.1 to 66.3) | 50.3 (46.8 to 53.0) | 91.3 (87.9 to 92.7) | 76.2 (67.2 to 80.1) |
| GBDT | 64.3 (61.5 to 67.1) | 86.7 (84.5 to 88.6) | 49.7 (46.0 to 53.4) | 63.0 (59.8 to 66.0) | 50.2 (46.1 to 53.4) | 91.9 (88.4 to 93.0) | 77.9 (68.6 to 80.9) |
| MLP | 57.7 (54.7 to 60.7) | 81.3 (78.9 to 83.7) | 44.2 (40.5 to 48.3) | 57.4 (54.3 to 60.5) | 46.1 (41.6 to 49.7) | 93.2 (89.4 to 94.1) | 81.4 (71.1 to 83.9) |
| Random forest | 61.9 (58.8 to 64.9) | 85.1 (82.9 to 87.2) | 43.6 (40.6 to 46.9) | 60.5 (57.2 to 63.7) | 44.8 (41.2 to 47.9) | 88.5 (85.6 to 90.8) | 68.9 (60.7 to 74.9) |
| gpt-oss-120b† | 59.2 (56.3 to 62.2) | 76.8 (74.2 to 79.5) | 50.0 (45.8 to 54.6) | 57.5 (54.4 to 60.7) | 46.9 (42.6 to 50.1) | 84.8 (81.2 to 85.9) | 58.8 (49.0 to 61.8) |
| **Data fusion** | | | | | | | |
| ClinicalBigBird | 66.5 (63.6 to 69.1) | 88.1 (86.0 to 89.9) | 52.2 (48.9 to 55.3) | 65.0 (62.0 to 67.9) | 51.5 (47.8 to 54.2) | 92.5 (89.5 to 93.9) | 79.5 (71.4 to 83.4) |
| ClinicalLongformer | 66.4 (63.4 to 69.1) | **91.0 (89.2 to 92.7)** | 55.5 (51.4 to 59.6) | 65.3 (62.1 to 68.1) | 55.8 (50.5 to 59.7) | 92.9 (90.0 to 94.5) | 80.6 (72.7 to 85.0) |
| gpt-oss-120b† | 65.4 (62.5 to 68.2) | 81.9 (79.6 to 84.0) | 59.6 (54.5 to 63.9) | 62.5 (59.2 to 65.6) | 56.0 (50.7 to 59.8) | 84.5 (81.3 to 86.0) | 57.9 (49.2 to 61.8) |
| **Feature fusion** | | | | | | | |
| AutoMM with BioMed-RoBERTa and attention | 66.8 (63.8 to 69.6) | 86.5 (84.4 to 88.5) | 56.8 (52.1 to 61.5) | 66.4 (63.3 to 69.2) | **57.5 (51.8 to 61.5)** | **95.7 (91.8 to 95.9)** | **88.3 (77.6 to 88.8)** |
| AutoMM with BioMed-RoBERTa | 63.2 (59.9 to 66.0) | 83.3 (80.9 to 85.6) | 53.4 (48.9 to 58.2) | 62.9 (59.5 to 65.6) | 52.6 (47.5 to 56.2) | 93.0 (89.4 to 94.3) | 80.9 (71.2 to 84.4) |
| **Decision fusion** | | | | | | | |
| Soft voting | 69.3 (66.4 to 72.1) | 90.9 (89.2 to 92.6) | 54.2 (50.8 to 57.5) | 67.8 (64.8 to 70.7) | 54.8 (51.1 to 57.7) | 92.7 (89.5 to 93.7) | 80.1 (71.4 to 82.8) |
| Class-aware voting | 68.3 (65.4 to 71.2) | 90.0 (88.3 to 91.8) | 53.7 (50.4 to 56.9) | 66.8 (63.9 to 69.9) | 54.3 (50.7 to 57.0) | 93.1 (89.8 to 94.0) | 81.2 (72.2 to 83.6) |
| Stacking | **69.5 (66.6 to 72.4)** | 88.3 (86.4 to 90.1) | 54.7 (50.7 to 58.3) | **68.2 (65.1 to 71.4)** | 56.6 (52.3 to 60.1) | 92.1 (89.0 to 93.6) | 78.4 (70.0 to 82.6) |

### S6 Calibration and uncertainty

All selected supervised pipelines had better calibration than InSilicoVA, with lower classwise ECE and lower Brier scores, computed from the full probability matrices (Table S9).

We estimated calibration on one retrospective test set and did not recalibrate or re-evaluate it after temporal or external shift.

Table S9: Classwise calibration metrics for the representative pipelines. Representative models: Baseline = InSilicoVA; unimodal narrative = BioMed-RoBERTa; unimodal structured = CatBoost; data fusion = ClinicalLongformer; feature fusion = AutoMM with BioMed-RoBERTa and attention.

| Pipeline | ECE | MCE | Brier |
|---|---|---|---|
| Baseline | 0.0350 | 0.9868 | 0.0386 |
| Unimodal narrative | 0.0147 | 0.9104 | 0.0271 |
| Unimodal structured | 0.0121 | 0.9207 | 0.0255 |
| Data fusion | 0.0149 | 0.8022 | 0.0244 |
| Feature fusion | 0.0252 | 0.8983 | 0.0284 |
| Soft voting | 0.0123 | 0.6049 | 0.0224 |
| Class-aware voting | 0.0125 | 0.8975 | 0.0224 |
| Stacking | 0.0097 | 0.8931 | 0.0226 |

Marginal confidence intervals were obtained by resampling the 1,000 held-out cases with replacement 1,000 times. They quantify sampling variability for a fixed fitted model. Model weights, training data, HPO, and the split remain fixed. They do not include training stochasticity or split uncertainty. The intervals for the representative pipelines are in Table 2, and those for every model in Table S8.

#### S6.1 Paired pipeline comparisons

All pipelines were scored on the same resampled test indices. Table S10 compares each multimodal pipeline, including the combined language-model arm, with the better unimodal representative for that metric. The 95% CIs were based on a paired bootstrap with 10,000 resamples. Positive values favor the multimodal pipeline. The paired intervals, rather than overlap of marginal intervals, support the metric-specific fusion claims.

Table S10: Paired bootstrap differences between each multimodal pipeline and the better unimodal representative for the stated metric. The reference is the structured-only pipeline throughout, except for the metric marked *, where the narrative-only pipeline was the more accurate of the two.

| Metric | Pipeline | Difference (95% CI) |
|---|---|---|
| Top-1 accuracy | gpt-oss-120b (combined) | +1.3 (−1.8 to +4.4) |
| | Data fusion | +2.3 (−0.4 to +5.1) |
| | Feature fusion | +2.7 (−0.2 to +5.6) |
| | Soft voting | +5.2 (+2.9 to +7.6) |
| | Class-aware voting | +4.2 (+2.0 to +6.5) |
| | Stacking | +5.4 (+3.1 to +7.7) |
| Balanced accuracy | gpt-oss-120b (combined) | +9.4 (+3.1 to +15.1) |
| | Data fusion | +5.3 (+0.3 to +10.5) |
| | Feature fusion | +6.6 (+1.7 to +11.5) |
| | Soft voting | +4.0 (−0.1 to +7.8) |
| | Class-aware voting | +3.5 (−0.6 to +7.3) |
| | Stacking | +4.4 (+0.3 to +8.5) |
| Macro F1 | gpt-oss-120b (combined) | +4.0 (−1.8 to +9.2) |
| | Data fusion | +3.8 (−2.0 to +9.4) |
| | Feature fusion | +5.5 (+0.6 to +10.3) |
| | Soft voting | +2.9 (−0.9 to +6.9) |
| | Class-aware voting | +2.3 (−1.4 to +6.4) |
| | Stacking | +4.6 (+0.5 to +8.9) |
| CSMF accuracy* | gpt-oss-120b (combined) | −7.7 (−10.8 to −4.9) |
| | Data fusion | +0.6 (−1.5 to +2.8) |
| | Feature fusion | +3.4 (−0.4 to +5.1) |
| | Soft voting | +0.4 (−2.0 to +2.4) |
| | Class-aware voting | +0.8 (−1.7 to +2.5) |
| | Stacking | −0.2 (−2.7 to +2.8) |

### S7 Cause-specific performance and error patterns

#### S7.1 Cause-specific accuracy by modality

The narrative pipeline was more accurate within noncommunicable causes and the structured pipeline within communicable causes, whether performance was weighted by deaths or equally across causes. For noncommunicable causes the narrative pipeline (BioMed-RoBERTa) reached 56.5% aggregate accuracy against 50.5% for the structured pipeline (CatBoost), and 53.6% against 46.7% when every cause is weighted equally. For communicable causes the structured pipeline reached 81.2% against 70.6%, and 35.1% against 23.8% cause-weighted. The largest narrative advantages were for liver cirrhosis, cerebrovascular disease, diabetes, and cardiac disease, and the largest structured advantages for pulmonary tuberculosis, meningitis and encephalitis, and pneumonia (Figure 1).

#### S7.2 Modality contribution and ensemble composition

Held-out top-1 accuracy was decomposed by modality for the supervised soft-voting ensemble and for the zero-shot language model, and a composition curve varied the ensemble's modality mix.

**Modality decomposition.** The players are the ten ensemble members, grouped into the five narrative PLMs and the five structured ML models, or, for the language model, its two inputs. The value of a coalition is its held-out top-1 accuracy: for the ensemble, soft voting restricted to those members, evaluated exactly over all $2^{10}$ subsets, and for the language model, the arm reading those inputs, with the empty coalition set to zero. Shapley values were computed per player and summed within modality, overall and within communicable and noncommunicable reference causes (Table S11).

Because every ordering of the players is equally likely, the size-zero term of a Shapley value is the player's own accuracy weighted by $1/k$. The value therefore splits into that solo part, which carries no information about the other modality, and a complementarity term $c_i$ collecting the gains from adding the player to nonempty coalitions:

$$\varphi_i = \underbrace{\tfrac{1}{k}\, v(\{i\})}_{\text{solo}_i} + c_i, \qquad \sum_i c_i = v(\text{all}) - \tfrac{1}{k}\sum_i v(\{i\}).$$

For two players this is $c_N = \frac{1}{2}[v(N{+}S) - v(S)]$ and $c_S = \frac{1}{2}[v(N{+}S) - v(N)]$: half the gain from adding that modality to the other.

The two decompositions form coalitions differently. A supervised coalition averages members that are fixed across coalitions, so adding one member does not change how another reads the record. Each language-model coalition is a separate run, and supplying both inputs changes how either one is read, which is a property of prompting a single model on joint input.

Table S11: Modality decomposition of held-out top-1 accuracy, in percentage points. Each Shapley value is split into its solo part and its complementarity term $c$, summed within modality: the four terms in a row sum to the combined pipeline's accuracy, and the two $c$ terms to its gain over the mean of that pipeline's individual members. A $c$ term is positive where the modality still adds accuracy once the other is present. Supervised values are computed exactly over all $2^{10}$ subsets of the ten ensemble members. The language-model values take the narrative and the structured responses as the two players. Intervals are 95% bootstrap intervals from 1,000 resamples of the held-out records, with the whole decomposition recomputed in each resample. Communicable and noncommunicable are Level 3 groupings of the reference cause and do not exhaust the held-out set.

| Reference cause group | Solo narrative | Solo structured | $c_{\text{narrative}}$ | $c_{\text{structured}}$ | Combined |
|---|---|---|---|---|---|
| *Supervised soft-voting ensemble, ten members* | | | | | |
| All causes ($n = 1{,}000$) | 31.4 | 31.4 | +3.70 (+1.36, +5.90) | +2.78 (+0.52, +5.07) | 69.3 |
| Communicable ($n = 330$) | 35.5 | 39.6 | −4.10 (−7.56, −0.82) | +7.83 (+4.71, +10.88) | 78.8 |
| Noncommunicable ($n = 382$) | 28.9 | 25.0 | +10.03 (+6.34, +14.05) | −2.12 (−5.83, +1.34) | 61.8 |
| *Zero-shot language model, two inputs* | | | | | |
| All causes ($n = 1{,}000$) | 30.7 | 29.6 | +3.10 (+1.80, +4.40) | +2.05 (+0.65, +3.40) | 65.4 |
| Communicable ($n = 330$) | 33.9 | 40.2 | −0.30 (−1.91, +1.50) | +5.91 (+3.70, +8.20) | 79.7 |
| Noncommunicable ($n = 382$) | 31.2 | 27.5 | +8.64 (+6.32, +11.02) | +4.97 (+2.75, +7.37) | 72.3 |

**Ensemble composition.** The composition analysis held the ensemble at five members and varied how many were narrative PLMs and how many were structured models, averaging over all compositions at each split (Figure S3). Individual-level performance followed an inverted U, peaking at the three-narrative and two-structured split. Both single-modality extremes fell three to four percentage points below that split on top-1 accuracy and were within 0.1 points of each other on macro F1, so neither modality alone reproduced the ensemble gain. Agreement with the population cause distribution varied little across compositions, and its highest value occurred with narrative models alone.

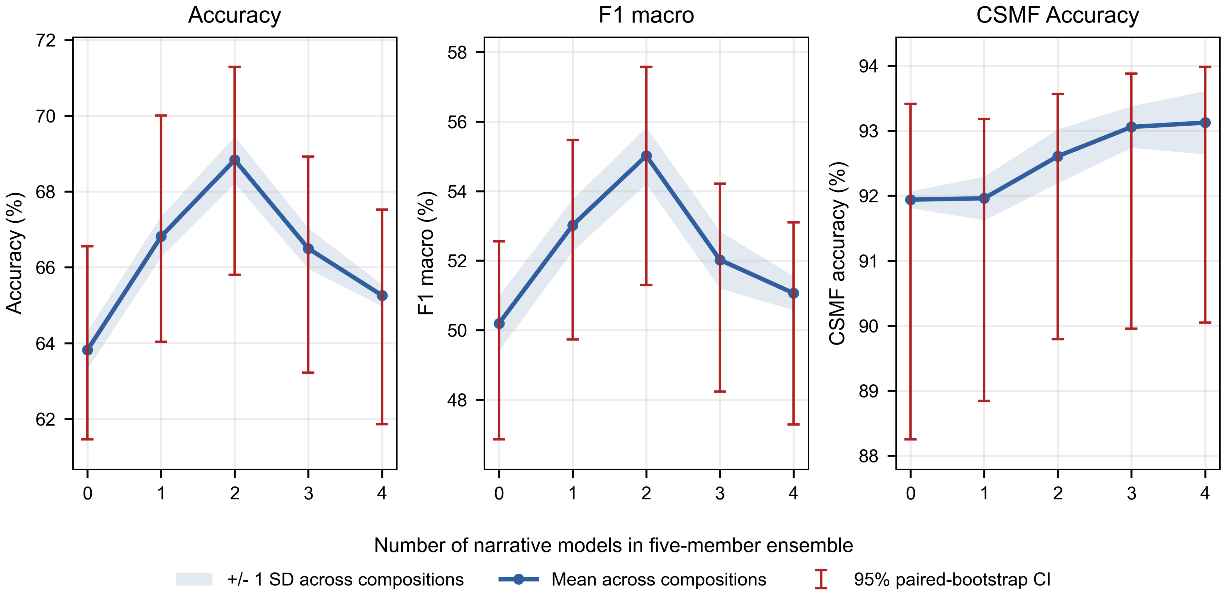

Figure S3: Soft-voting performance by modality composition. Each point is the mean across all five-member ensembles with the stated number of narrative and structured base models. Bands span one standard deviation across compositions, and error bars are 95% paired-bootstrap confidence intervals.

#### S7.3 Fusion gains by cause prevalence

The fusion levels gained on different causes, which accounts for their different metric profiles. Table S12 aggregates the cause-specific accuracies shown in Figure 1: causes are grouped by how many held-out deaths they carry, and each pipeline is compared with whichever unimodal representative is more accurate in that band. Decision fusion gains on the commonest causes and little or nothing on the rarest. Data-level and feature-level fusion do the reverse. Top-1 accuracy is dominated by the commonest causes, whereas macro F1 and balanced accuracy weight every cause equally, so the same pattern produces the different metric rankings reported in Table 2. Stratified by rarity, feature fusion's advantage across causes is concentrated in the rarest causes ($n < 10$): restricted to the 16 causes carrying at least 10 held-out deaths (per-cause F1 averaged over those causes), the macro F1 values of feature fusion and the three decision-fusion pipelines fall within 1.6 percentage points of one another (59.4 to 61.0), and every paired difference between feature fusion and a decision-fusion pipeline has a 95% interval spanning zero: $-1.6$ ($-5.3$ to $+1.9$) against stacking, $-1.3$ ($-5.4$ to $+2.4$) against soft voting, and $-0.3$ ($-4.3$ to $+3.3$) against class-aware voting.

Table S12: Cause-specific recall gain over the better unimodal representative, by cause prevalence (mean percentage points). Narrative = BioMed-RoBERTa; structured = CatBoost; data fusion = ClinicalLongformer; feature fusion = AutoMM with BioMed-RoBERTa and cross-modal attention. Decision-fusion pipelines combine the same ten base models.

| Pipeline | Common causes (60+ deaths) | Intermediate causes (20–59 deaths) | Rare causes (<20 deaths) |
|---|---|---|---|
| Data fusion | 0.0 | 1.9 | 8.5 |
| Feature fusion | 0.7 | 4.6 | 8.7 |
| Soft voting | 4.5 | 4.5 | -0.9 |
| Class-aware voting | 3.3 | 4.0 | -0.9 |
| Stacking | 5.1 | 1.9 | 2.5 |

**A case reading of the meningitis deaths.** Meningitis is the cause on which the fusion levels separate most sharply, and its five held-out deaths show why. The two modalities ranked the cause highly on *different* deaths: taking the median rank assigned to the correct cause across the five models of each modality, the structured models placed meningitis second or third on two deaths and sixth or lower on the other three, while on exactly those three the narrative models placed it third or fourth. The competing label was HIV/AIDS, cerebrovascular disease, or an indeterminate assignment. Feature fusion ranked the correct cause first or second on all five deaths and data fusion on three, whereas soft voting and class-aware voting ranked it fourth to sixth throughout and the meta-learner ranked it worse than the stronger modality on every one of the five, consistent with a second stage fitted on out-of-fold predictions in which the cause is almost absent. Five deaths make this an illustration rather than a test, and it cannot be read as evidence that fusion recovers more of the underlying cause of death: the reference was assigned by physicians who read the narrative, so a pipeline that reads it too is by construction closer to the coder's information set.

#### S7.4 Error patterns and mechanisms

**Partition by unimodal correctness.** The main error partition uses the same representatives as Table 2: BioMed-RoBERTa, CatBoost, and AutoMM with BioMed-RoBERTa plus cross-modal attention. Figure S4 extends the partition to comparisons with InSilicoVA.

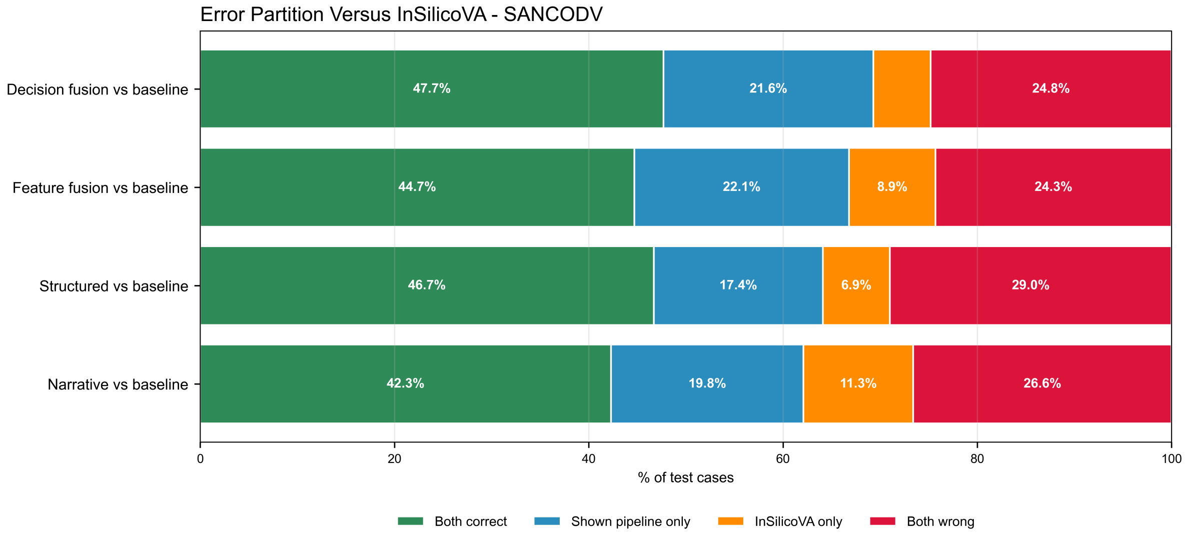

Figure S4: Pairwise error partitions versus InSilicoVA for the representative narrative, structured, feature-fusion, and soft-voting pipelines. Narrative text = BioMed-RoBERTa; structured = CatBoost; feature fusion = AutoMM with BioMed-RoBERTa and cross-modal attention. Held-out test set ($n$ = 1,000).

**Deaths no supervised representative classified correctly.** Errors shared across pipelines concentrated in the least specific categories. Of the 176 held-out deaths (17.6%) that all seven supervised representatives misclassified, 37 were coded indeterminate and 51 to one of the three residual "other and unspecified" categories — half the set, from four categories carrying a quarter of the held-out deaths. All 13 other and unspecified infection deaths were missed by every pipeline, as were 52.8% of other and unspecified noncommunicable, 38.5% of other and unspecified external, and 23.7% of indeterminate deaths. This set is not the one in the next paragraph, which is defined over the ten decision-fusion base models rather than the seven representatives: 127 deaths belong to both, 49 were missed by every representative although some base model assigned them correctly, and 16 were missed by every base model but recovered by a fusion pipeline.

**Deaths missed by every base model.** Table S13 partitions the held-out set by how many of the ten soft-voting members classified each death correctly. On the 143 deaths that all ten missed, none of the three decision-level combiners recovered a case, whereas feature fusion and data fusion — independently trained models that are not members — recovered a few. This shared failure set is an observed property of the three combiners evaluated here rather than an arithmetic constraint: soft voting averages the members' full probability vectors rather than their votes, so a cause that several members rank second can still come first after averaging. On these deaths it never did. The correct cause was rarely absent from the members' candidate sets — at least one member ranked it within its top three in 124 of the 143 — but averaging demoted it to a median rank of four. The asymmetry is in the probability mass rather than the ranking: the correct cause held a mean averaged probability of 0.061 against 0.523 for the cause that won. Averaging preserves a confident consensus, and on these deaths the members agreed confidently on the wrong cause.

**Rare-cause assignment across learning regimes.** The seven causes carrying fewer than 20 held-out deaths (72 deaths) separate the pipelines by how they were fitted rather than by modality or fusion level. Every supervised pipeline assigned fewer deaths to these causes than the reference standard contains, at 0.49 to 0.69 times their reference share, the narrative and structured representatives identically at 0.56. Every zero-shot language-model arm assigned more, at 1.08 to 1.74 times, as did InSilicoVA at 1.40. The direction is unchanged when the rare set is widened

Table S13: Top-1 accuracy (%) of each fusion pipeline, by how many of the ten soft-voting members classified the death correctly. Data fusion = ClinicalLongformer and feature fusion = AutoMM with BioMed-RoBERTa and cross-modal attention are trained independently and are not members of the ensemble. The three decision-level combiners are functions of the members.

| Pipeline | All 10 wrong | 1–4 correct | 5 correct | 6 or more correct |
|---|---|---|---|---|
| Held-out deaths (n) | 143 | 167 | 105 | 585 |
| Data fusion (not a member) | 4.2 | 29.3 | 61.9 | 93.0 |
| Feature fusion (not a member) | 7.0 | 35.9 | 58.1 | 91.8 |
| Soft voting | 0.0 | 18.0 | 76.2 | 99.7 |
| Class-aware voting | 0.0 | 17.4 | 70.5 | 99.1 |
| Stacking | 0.0 | 24.0 | 70.5 | 99.3 |

to causes below 30 or below 40 held-out deaths. Recall on these deaths split the same way in each comparison at matched input: 59.7% for the language model reading the narrative against 33.3% for BioMed-RoBERTa, 43.1% against 36.1% for CatBoost on the structured responses, and 47.2% against 44.4% for ClinicalLongformer on both.

#### S7.5 Confusion matrices

Figures S5–S9 show row-normalized confusion matrices for each representative pipeline, followed by the three decision-fusion pipelines and the three LLM arms (Figures S13–S15).

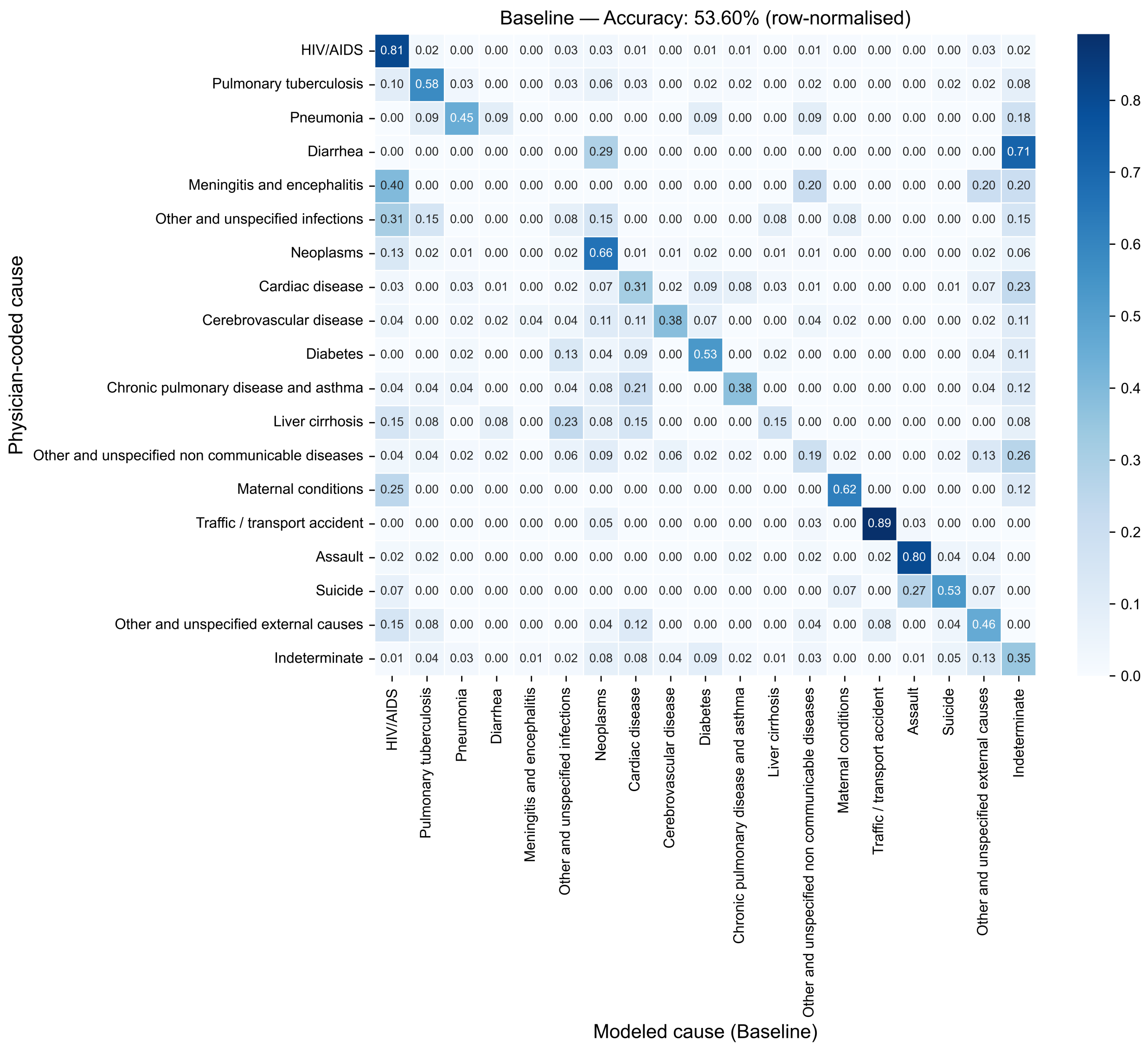

Model detail: Baseline = InSilicoVA. Values are row-normalized within physician-coded cause.

Figure S5: Row-normalized confusion matrix for InSilicoVA, the operational comparator.

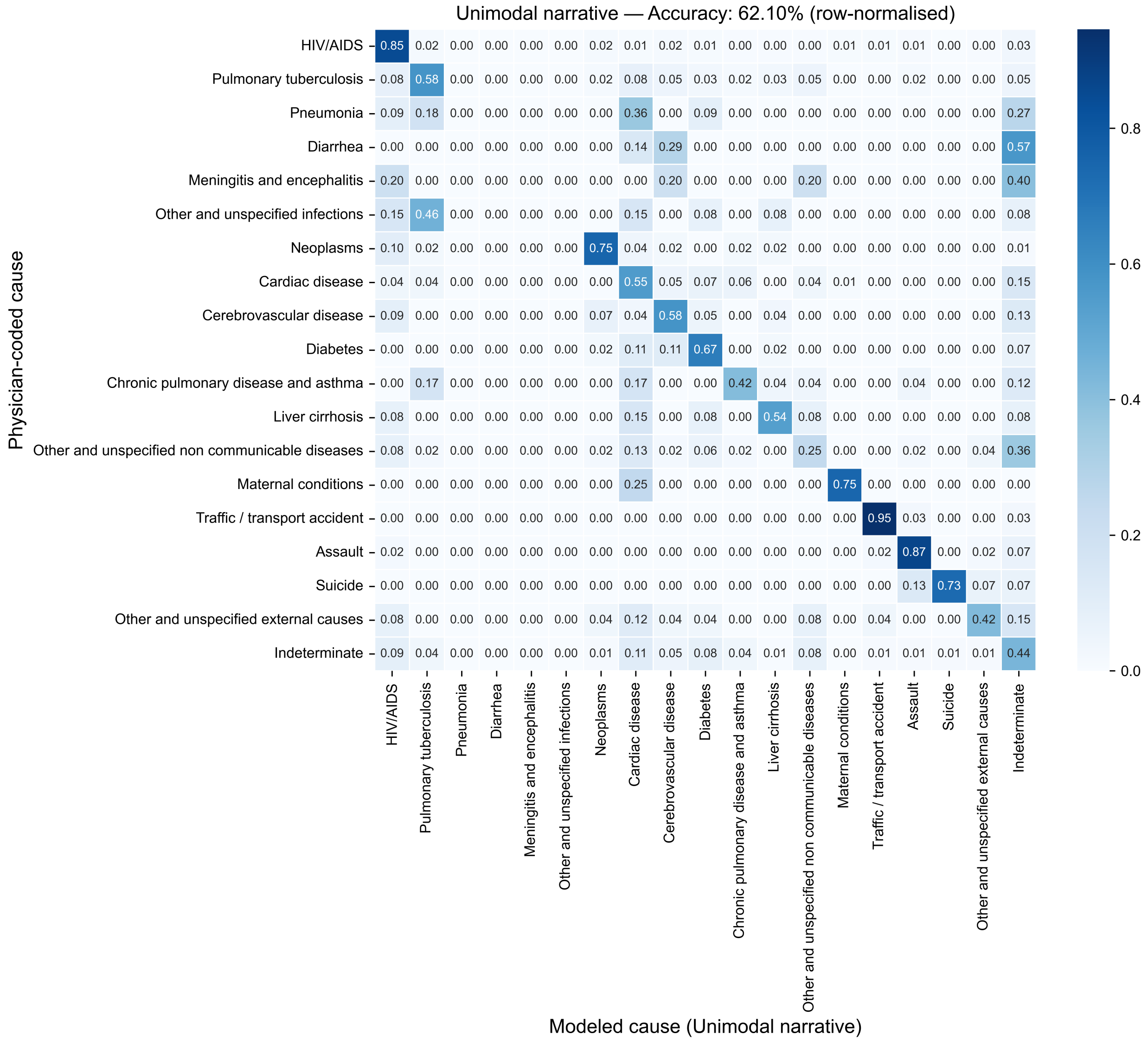

Model detail: Unimodal narrative = BioMed-RoBERTa. Values are row-normalized within physician-coded cause.

Figure S6: Row-normalized confusion matrix for BioMed-RoBERTa, the representative narrative model.

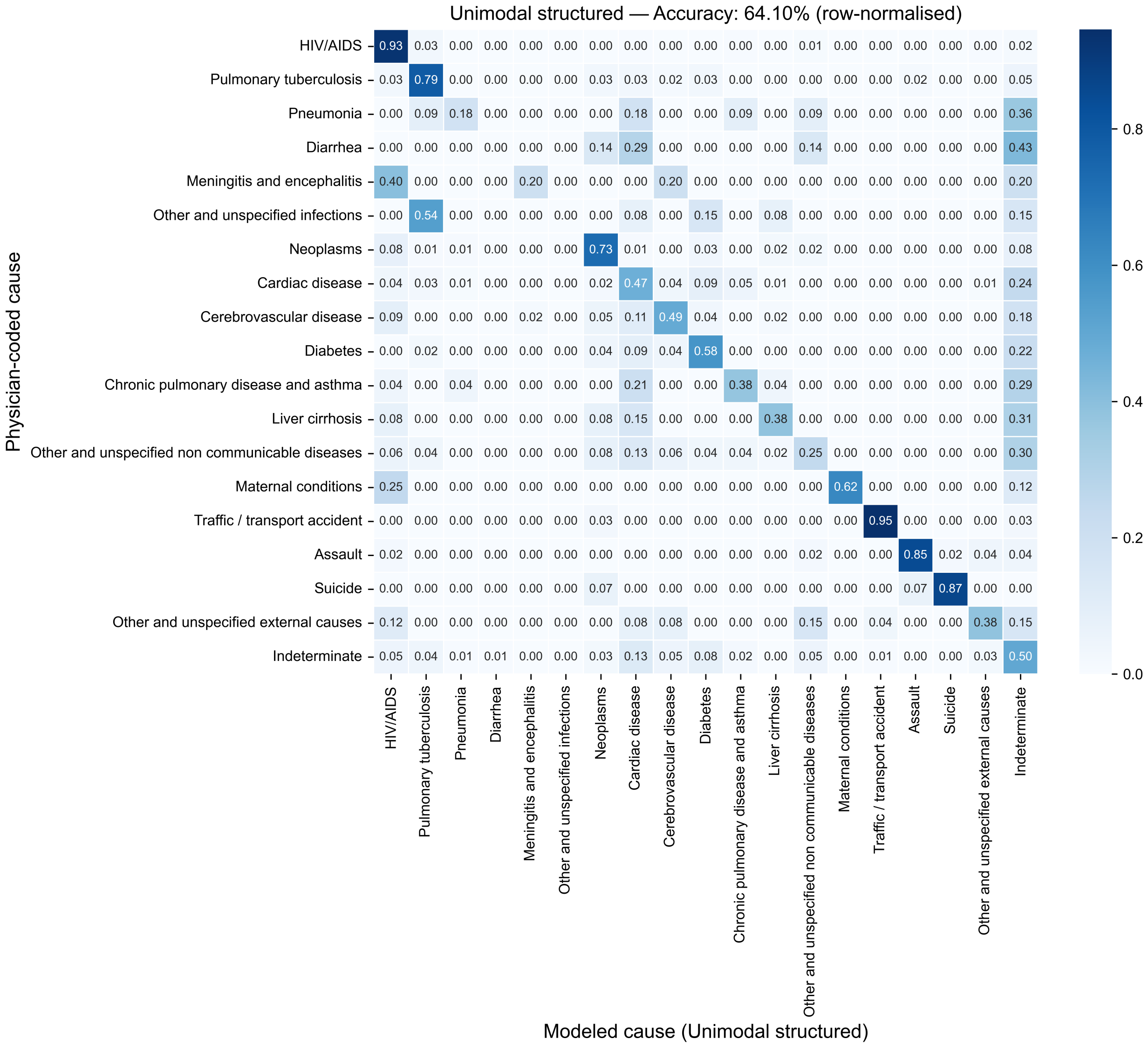

Model detail: Unimodal structured = CatBoost. Values are row-normalized within physician-coded cause.

Figure S7: Row-normalized confusion matrix for CatBoost, the representative structured model.

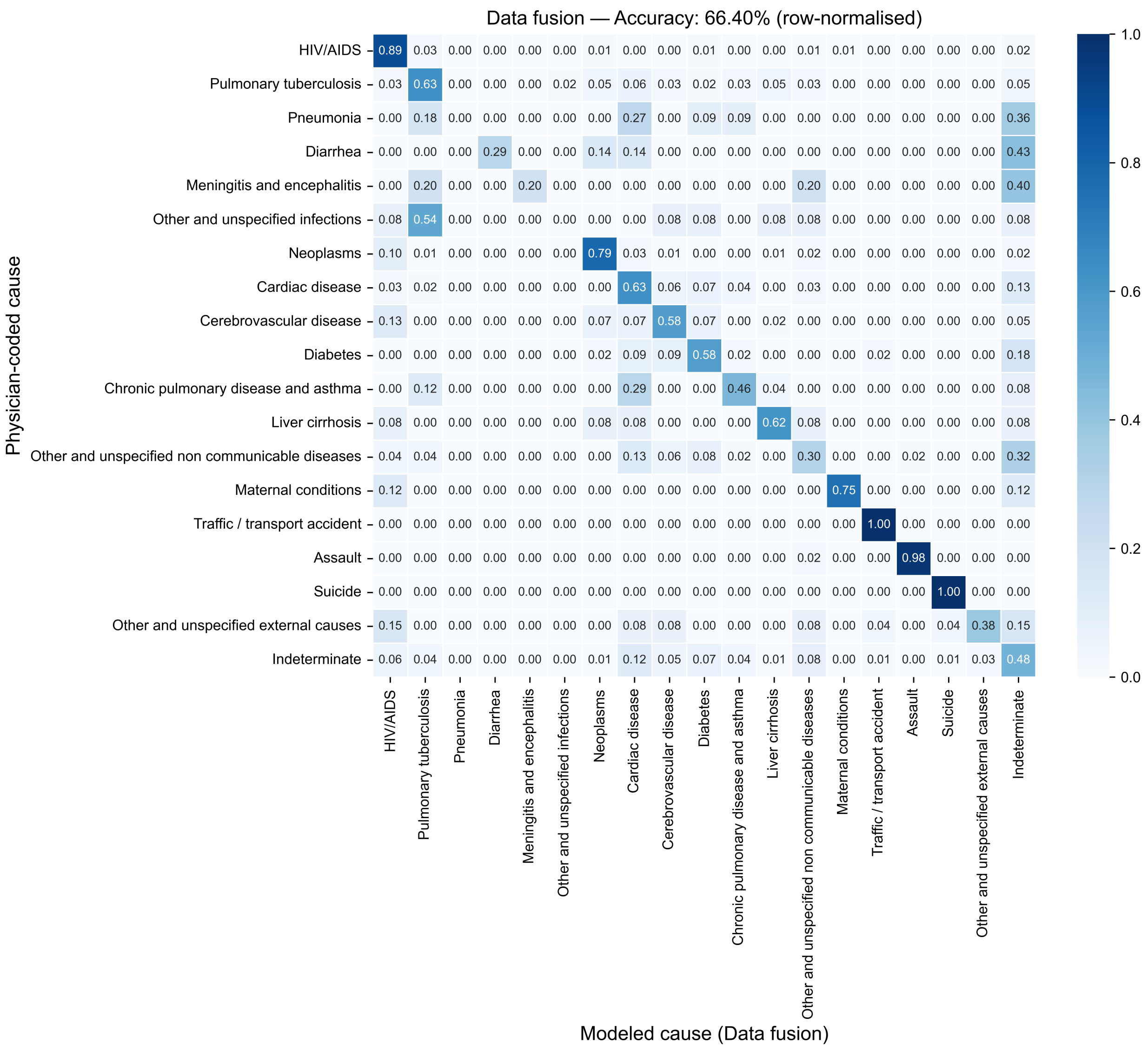

Model detail: Data fusion = ClinicalLongformer. Values are row-normalized within physician-coded cause.

Figure S8: Row-normalized confusion matrix for ClinicalLongformer, the representative data-fusion model.

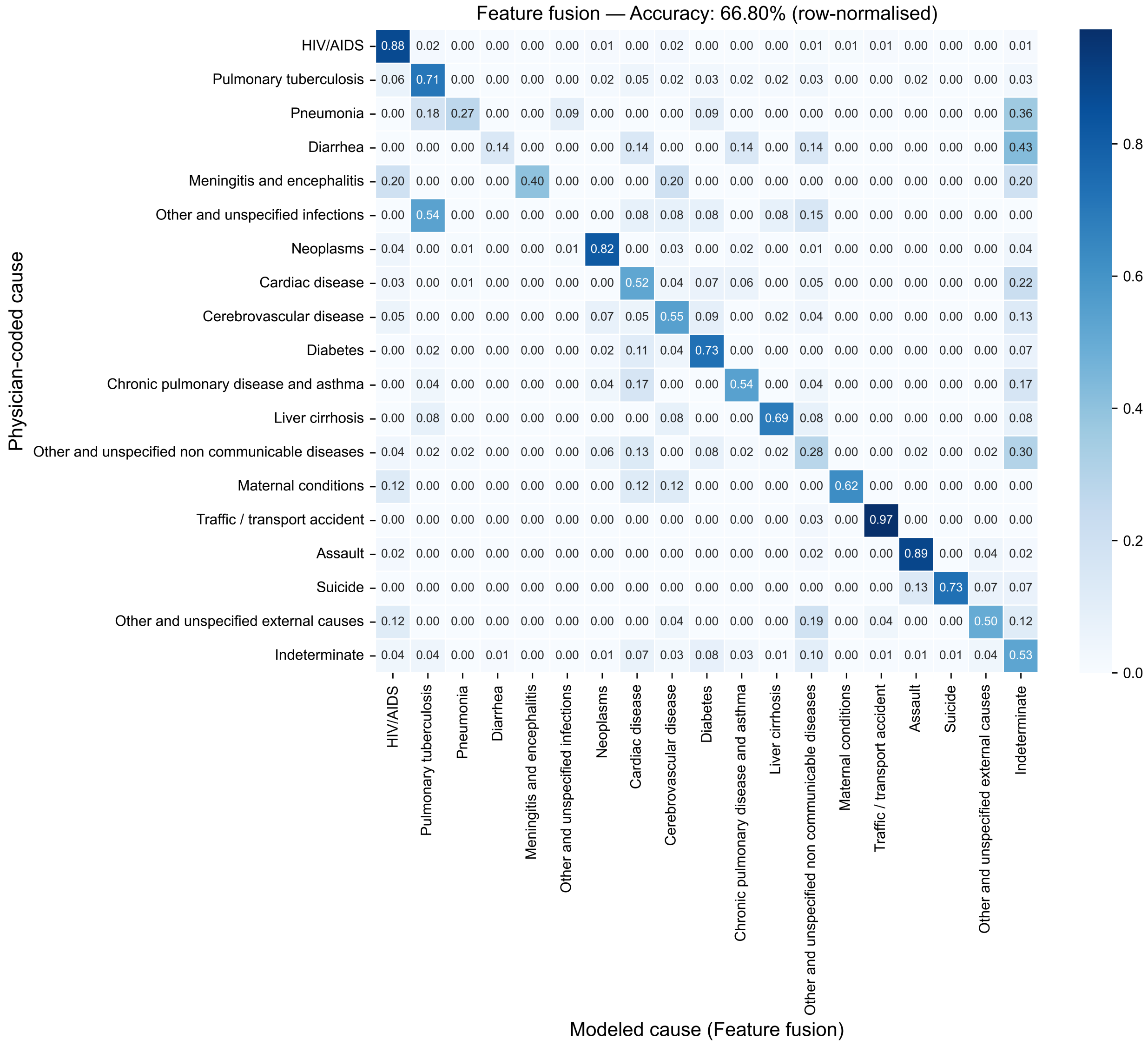

Model detail: Feature fusion = AutoMM with BioMed-RoBERTa and attention. Values are row-normalized within physician-coded cause.

Figure S9: Row-normalized confusion matrix for AutoMM with BioMed-RoBERTa and cross-modal attention, the representative feature-fusion model.

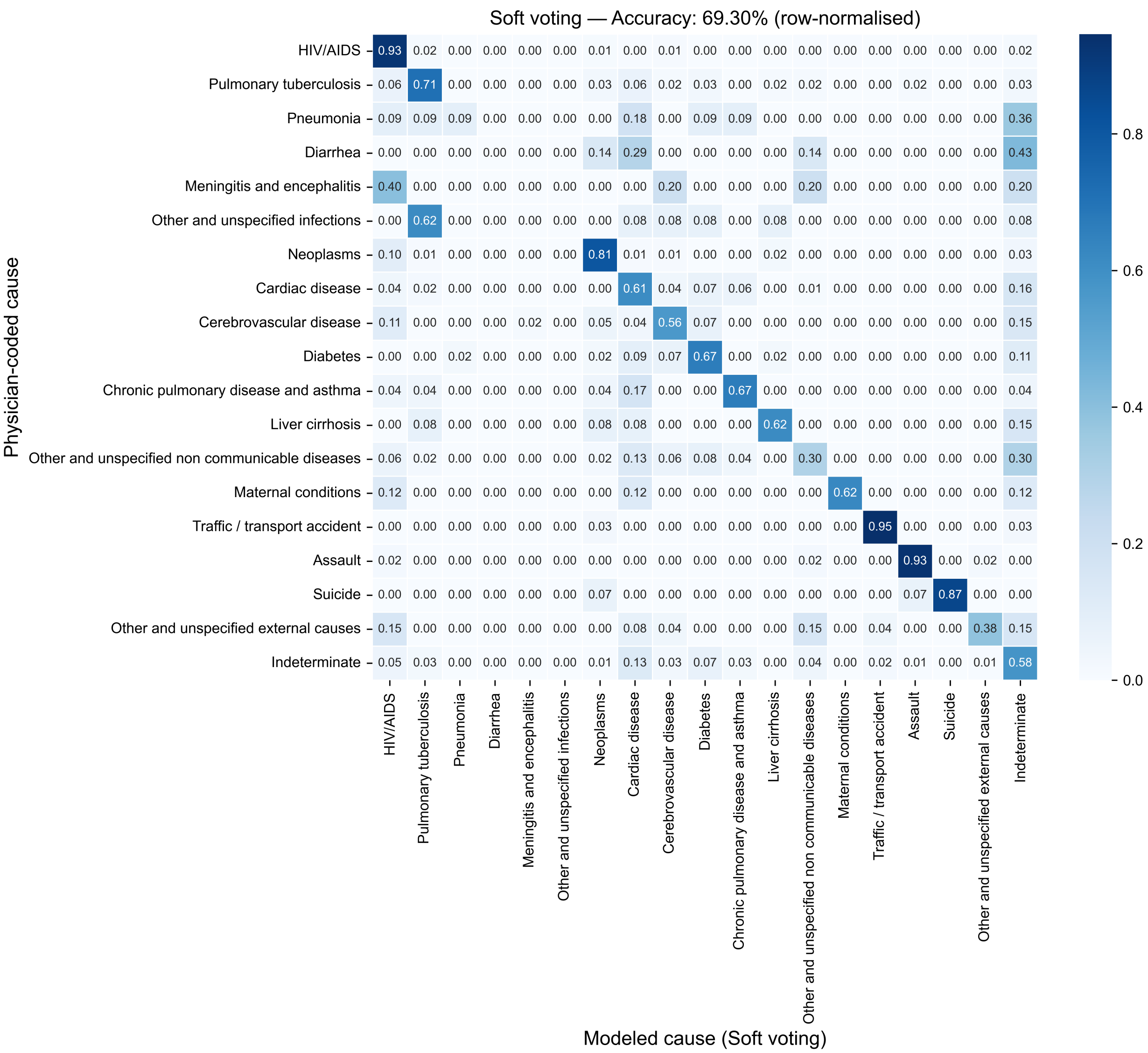

Model detail: Soft voting = Soft voting. Values are row-normalized within physician-coded cause.

Figure S10: Row-normalized confusion matrix for soft voting.

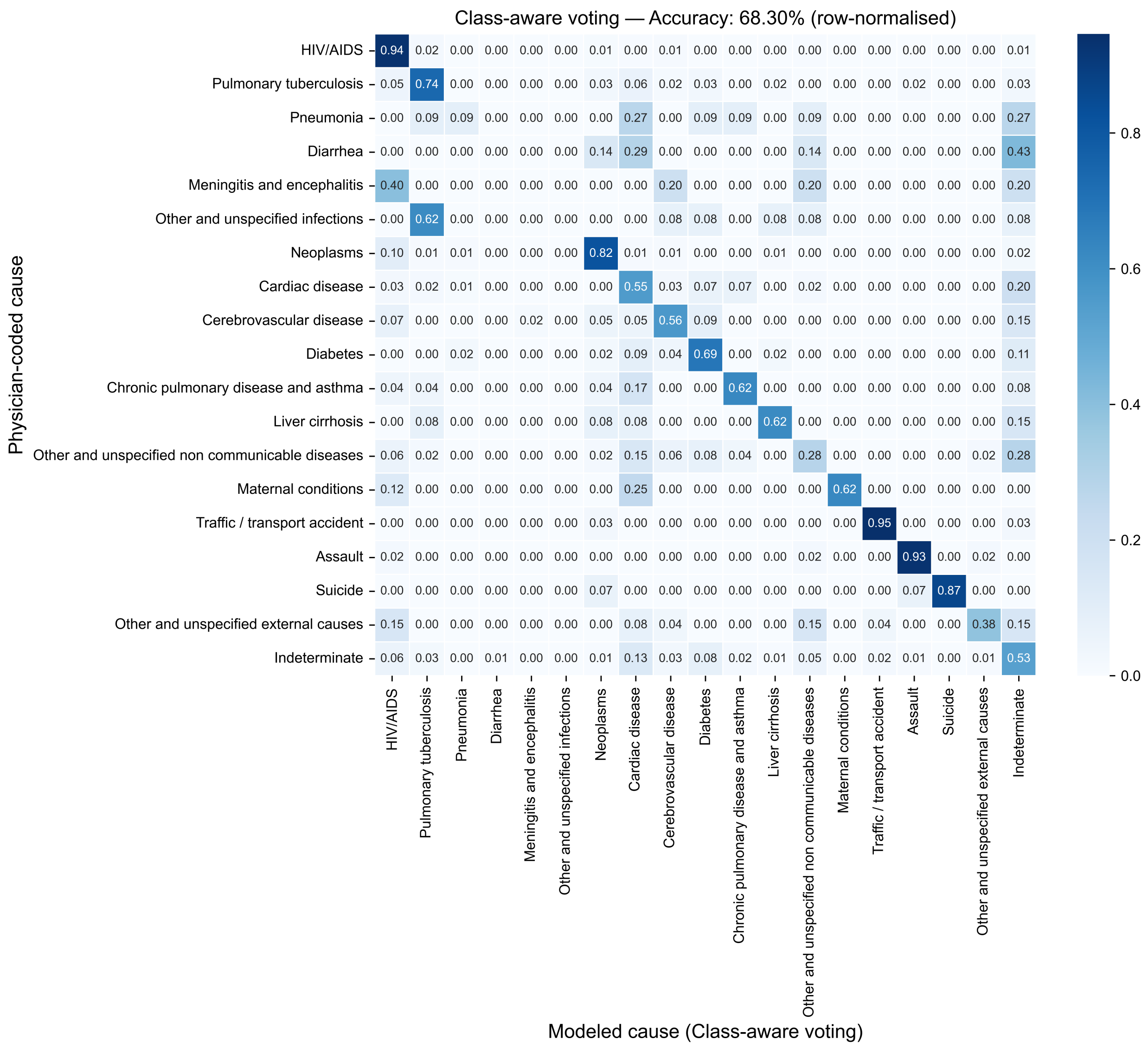

Model detail: Class-aware voting = Class-aware voting. Values are row-normalized within physician-coded cause.

Figure S11: Row-normalized confusion matrix for class-aware voting.

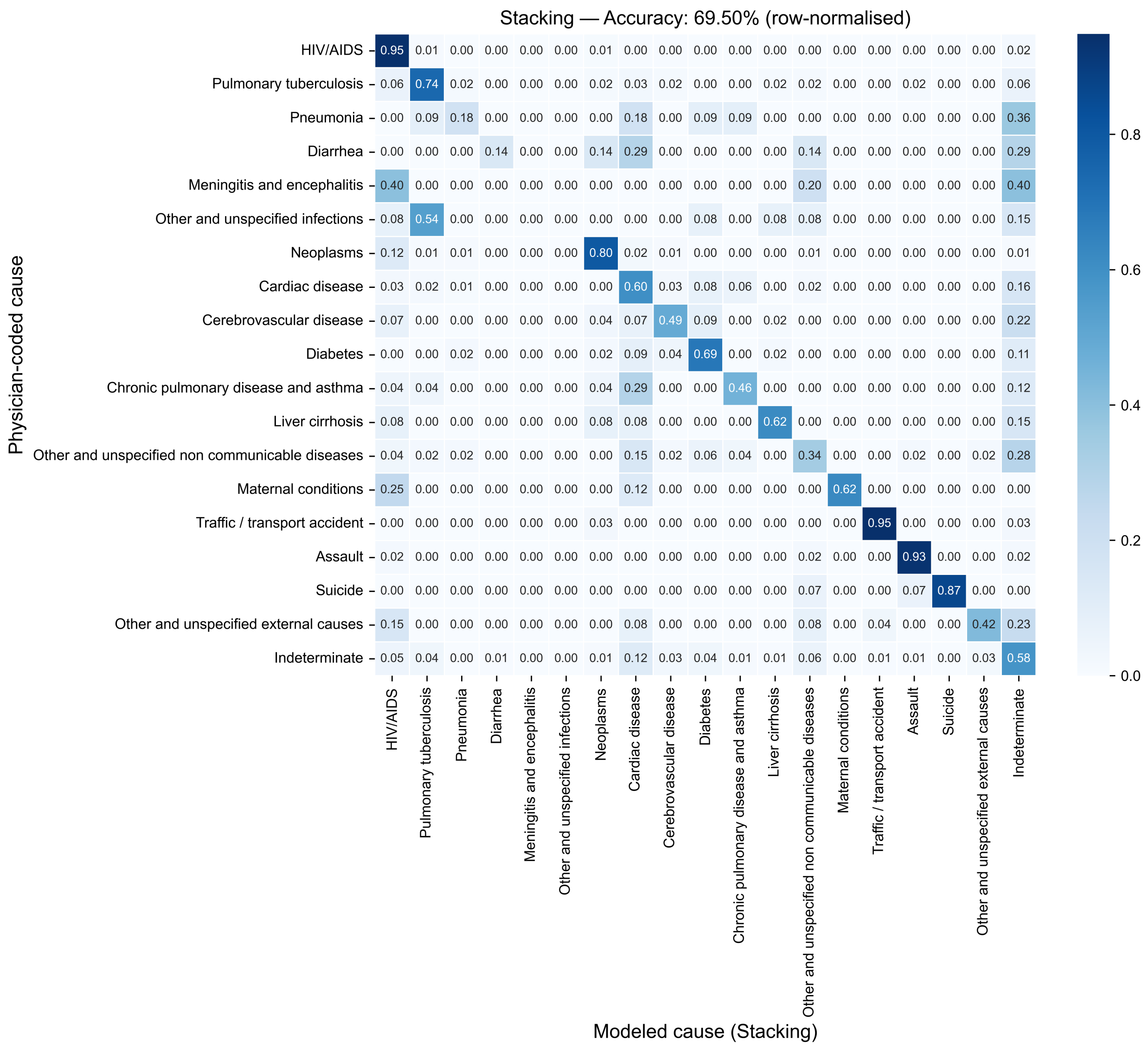

Model detail: Stacking = Stacking. Values are row-normalized within physician-coded cause.

Figure S12: Row-normalized confusion matrix for stacking.

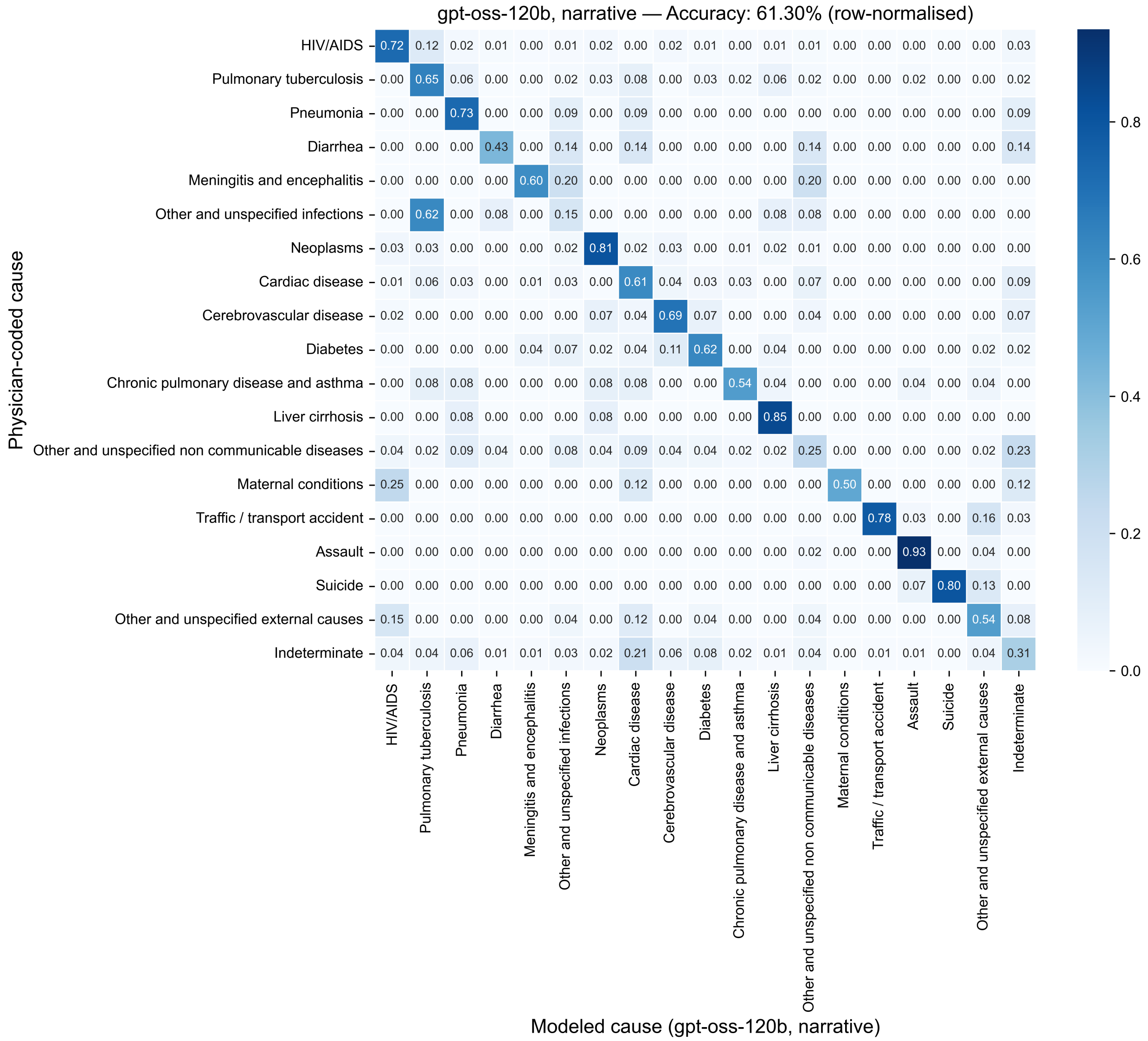

Model detail: gpt-oss-120b, narrative = gpt-oss-120b (narrative). Values are row-normalized within physician-coded cause.

Figure S13: Row-normalized confusion matrix for the zero-shot `gpt-oss-120b` arm reading the narrative alone (Section S4.4).

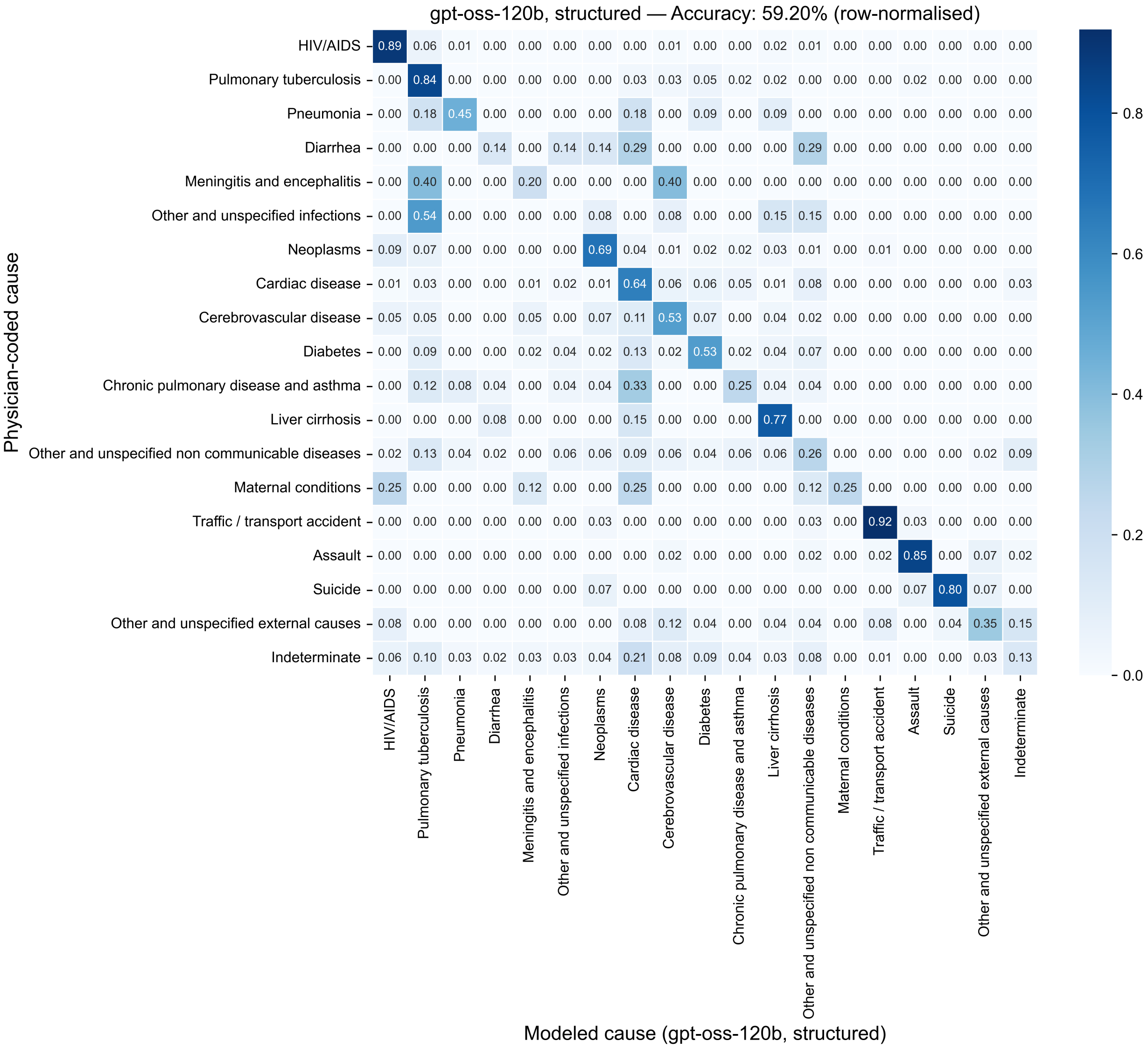

Model detail: gpt-oss-120b, structured = gpt-oss-120b (structured). Values are row-normalized within physician-coded cause.

Figure S14: Row-normalized confusion matrix for the zero-shot `gpt-oss-120b` arm reading the structured responses alone (Section S4.4).

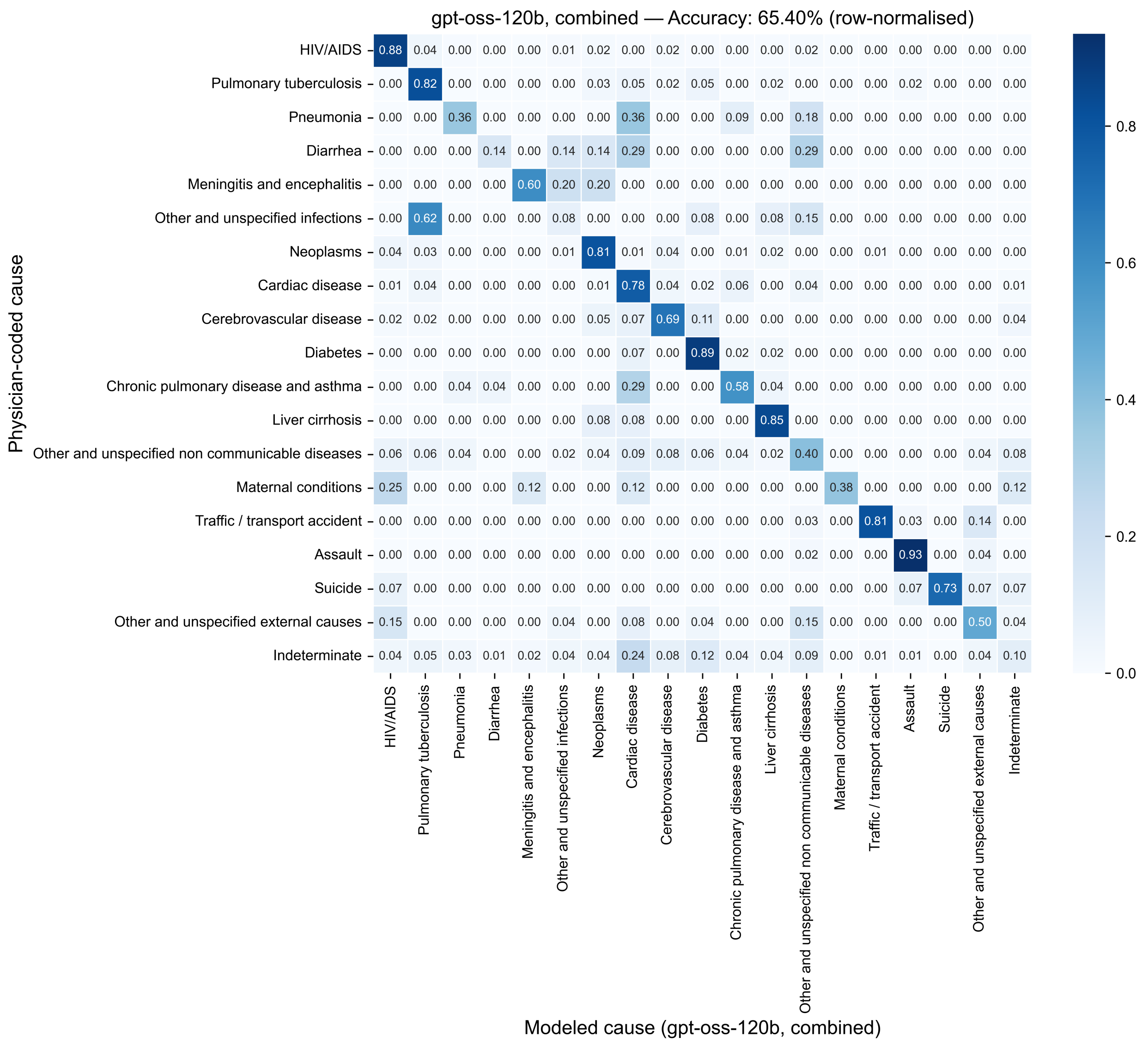

Model detail: gpt-oss-120b, combined = gpt-oss-120b (combined). Values are row-normalized within physician-coded cause.

Figure S15: Row-normalized confusion matrix for the zero-shot `gpt-oss-120b` arm reading the narrative and the structured responses together (Section S4.4). Compared with the narrative arm, assignments to the residual category fall from 89 to 25 of 1,000 records against a reference share of 156.

### S8 Robustness and sensitivity

Sensitivity analyses of the InSilicoVA specification and MCMC settings are reported in Section S4.3.1.

#### S8.1 COD grouping sensitivity

Comparing the endpoints, top-1 accuracy and macro F1 were higher at Level 3 than at Level 1 for every pipeline (Figure S16). CSMF accuracy was also higher at Level 3 for every pipeline except the primary feature-fusion model. At Level 3, every supervised pipeline assigned fewer deaths to indeterminate cause and more to noncommunicable disease than the reference standard, consistent with the models assigning a specific cause where physician review remained indeterminate.

The feature-fusion exception was specific to the fitted model. Aggregating its 19-cause predictions into the six broader groups produced nearly the same individual-level accuracy as training directly on six causes but substantially closer agreement with the population cause distribution. Replacing the narrative backbone reversed the CSMF pattern while leaving top-1 accuracy essentially unchanged. The decline in CSMF accuracy therefore should not be interpreted as a general consequence of training on broader cause groups.

The data-fusion searches completed fewer hyperparameter trials at the finer and broader grouping levels than at the primary level (15 at Level 1 and Level 3 against 29 at the primary level), so some across-level variation may reflect tuning effort. The 19-cause task remained the prespecified primary analysis. Chance-corrected CSMF accuracy was not compared across grouping levels because its chance constant does not vary with the number of causes.

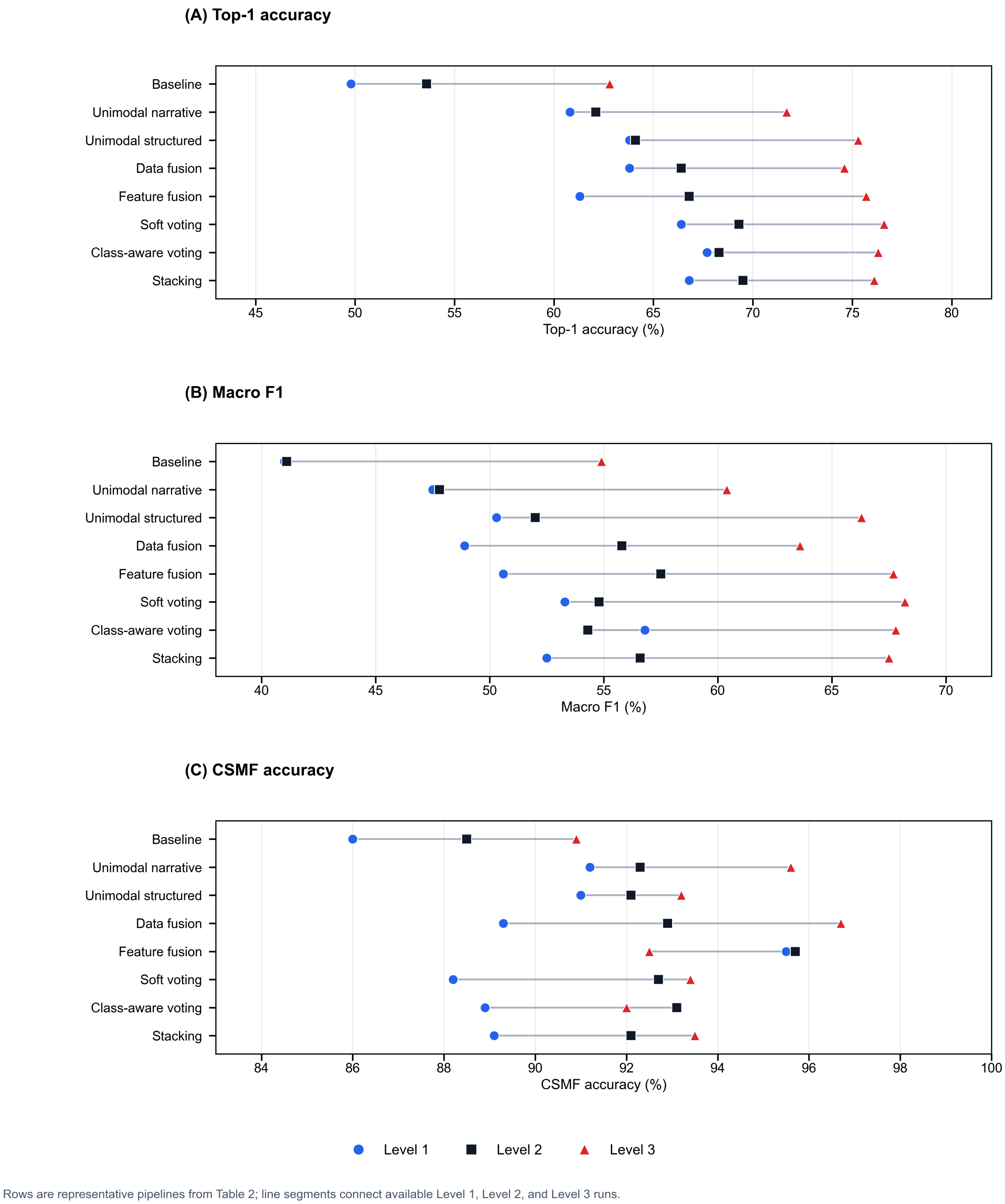

Figure S16: Held-out top-1 accuracy, macro F1, and CSMF accuracy by cause-grouping level (Level 1, 29 causes; Level 2, 19 causes, the primary analysis; Level 3, six causes). Representative model per pipeline: baseline = InSilicoVA; unimodal narrative = BioMed-RoBERTa; unimodal structured = CatBoost; data fusion = ClinicalLongformer; feature fusion = AutoMM with BioMed-RoBERTa and cross-modal attention.

#### S8.2 Feature-fusion architecture and backbone sensitivity

With the narrative backbone, training partition, and training budget held fixed, cross-modal attention produced higher CSMF accuracy than MLP fusion under either structured encoder. The categorical-embedding and cross-modal-attention configuration was also strongest on the individual-level metrics. Replacing BioMed-RoBERTa with BiomedBERT, the runner-up on training-partition cross-validated macro F1, produced small differences whose paired intervals all included zero. The two backbone configurations assigned the same cause to 80.5% of held-out deaths (Table S14).

Table S14: Feature-fusion sensitivity to architecture and narrative backbone. Panel A holds the BioMed-RoBERTa backbone fixed and varies the structured encoder and fusion module. Panel B holds the primary architecture fixed and varies the narrative backbone.

*A. Architecture sensitivity*

| Structured encoder | Fusion module | Top-1 accuracy | Balanced accuracy | Macro F1 | CSMF accuracy |
|---|---|---|---|---|---|
| Categorical and numerical embeddings | Cross-modal attention* | 66.8 | 56.8 | 57.5 | 95.7 |
| Feature-tokenizer transformer | Cross-modal attention | 61.8 | 54.1 | 52.9 | 95.6 |
| Categorical and numerical embeddings | MLP | 62.9 | 51.5 | 52.1 | 94.2 |
| Feature-tokenizer transformer | MLP | 63.2 | 53.4 | 52.6 | 93.0 |

*B. Narrative-backbone sensitivity*

| Metric | BioMed-RoBERTa | BiomedBERT | Difference (95% CI) |
|---|---|---|---|
| Top-1 accuracy | 66.8 | 65.8 | +1.0 (−1.1 to +3.1) |
| Balanced accuracy | 56.8 | 55.1 | +1.7 (−2.1 to +5.6) |
| Macro F1 | 57.5 | 56.5 | +1.0 (−2.7 to +5.1) |
| CSMF accuracy | 95.7 | 95.1 | +0.6 (−2.2 to +3.0) |

*Note.* Metrics are percentages. All configurations used the same training partition, AutoMM preset, and 24-hour training budget without configuration-specific hyperparameter search. The asterisk marks the primary architecture. Panel B differences are BioMed-RoBERTa minus BiomedBERT, estimated by paired bootstrap of the held-out set with 10,000 resamples.

#### S8.3 Leave-one-province-out

The nine provinces differ markedly in both size and cause profile, which is what makes them useful as held-out domains. Held-out sample size ranged from 185 records in Western Cape to 1,026 in North West. HIV/AIDS ranged from 13.5% of deaths in Western Cape to 26.2% in KwaZulu-Natal, and indeterminate cause from 9.7% to 22.0%.

For each province, model fitting excluded that province and evaluation used only its records. Hyperparameters were frozen at primary-analysis values. Those values had been chosen using training data spanning all provinces. Consequently, this is a domain-generalization sensitivity analysis using globally selected hyperparameters, not fully nested province-isolated HPO and not external validation.

Nine-province outputs are complete for all eight pipelines reported in Table 2. The feature-fusion folds were fitted under a shorter per-fold training budget (three hours) than the primary analysis. The budget was identical across the nine folds, so comparisons within this analysis are internally consistent. LOPO values are therefore not directly comparable with the held-out primary results, which were fitted under a longer budget.

Every supervised pipeline exceeded InSilicoVA for top-1 accuracy in all nine held-out provinces (Table S15) (Figure S17). Of the 63 pipeline–province comparisons for each metric, one fell below InSilicoVA on macro F1 (BioMed-RoBERTa in Western Cape) and four on CSMF accuracy (feature fusion in Limpopo, soft voting, class-aware voting, and stacking in North West, the province where InSilicoVA's CSMF accuracy was highest). Province-to-province spread tracked the narrative: the narrative-only, data-fusion and feature-fusion pipelines varied most in top-1 accuracy, over ranges of 11.9 to 14.1 points, against 6.4 to 6.5 points for CatBoost and soft voting. Western Cape, the smallest held-out province at 185 records against 309 to 1,026 elsewhere, was the weakest fold for nearly every pipeline.

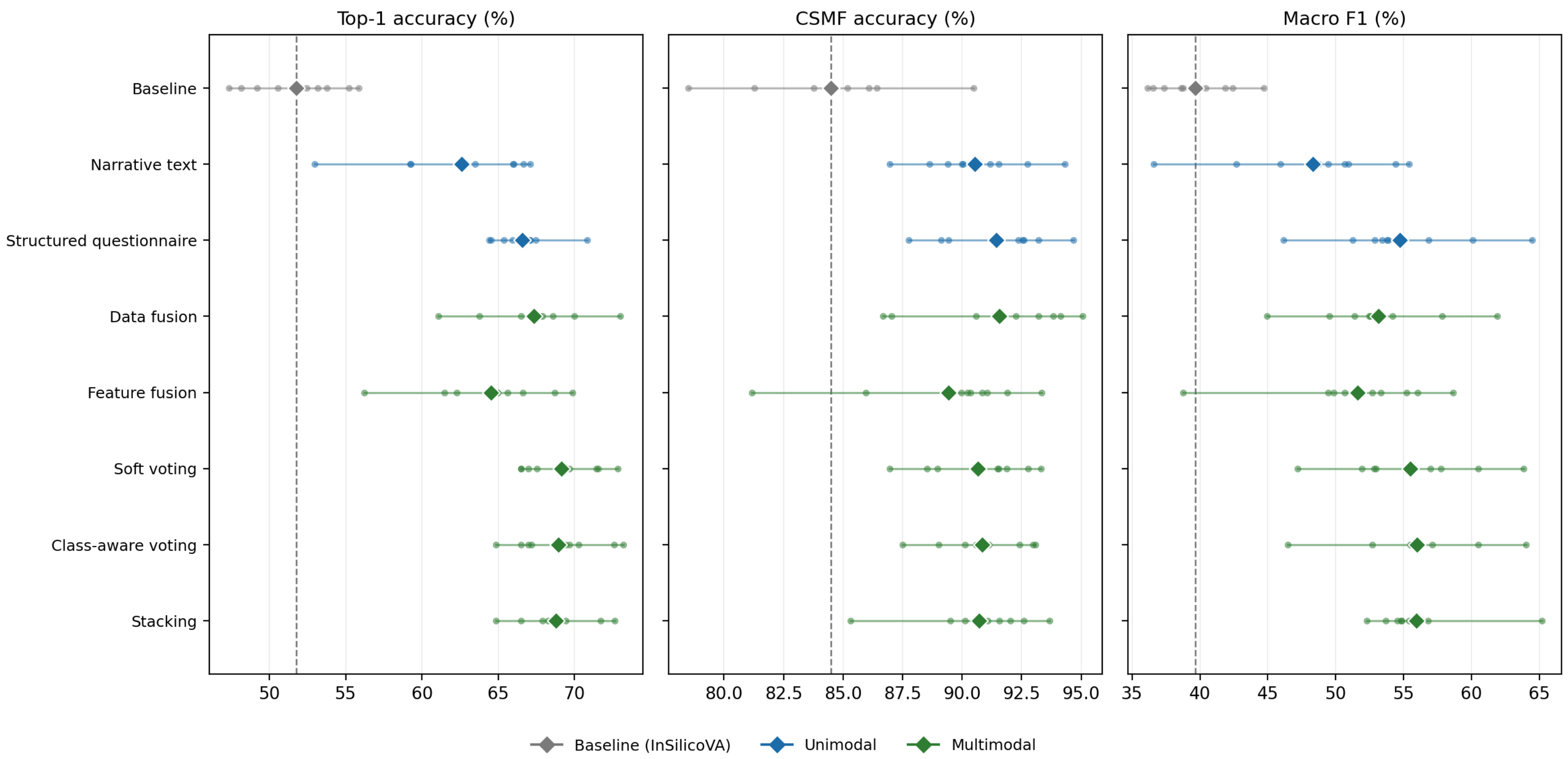

Figure S17: LOPO performance across nine held-out provinces. Circles are province-specific values, diamonds are unweighted means, and horizontal lines span observed ranges. Representative model per pipeline: comparator = InSilicoVA; narrative = BioMed-RoBERTa; structured = CatBoost; data fusion = ClinicalLongformer; feature fusion = AutoMM with BioMed-RoBERTa and cross-modal attention.

Table S15: LOPO evaluation. Each model was refit on eight provinces and evaluated on the ninth with globally selected hyperparameters frozen.

| Pipeline | EC (556) | FS (617) | GT (427) | KZN (841) | LM (309) | MP (567) | NC (471) | NW (1,026) | WC (185) | Mean | Range |
|---|---|---|---|---|---|---|---|---|---|---|---|
| **Top-1 accuracy (%)** | | | | | | | | | | | |
| *Operational comparator* | | | | | | | | | | | |
| Baseline | 55.2 | 47.3 | 53.2 | 53.7 | 49.2 | 48.1 | 50.5 | 55.8 | 52.4 | 51.7 | 8.5 |
| *Unimodal* | | | | | | | | | | | |
| Narrative text | 66.0 | 66.0 | 59.3 | 62.8 | 59.2 | 63.5 | 67.1 | 66.7 | 53.0 | 62.6 | 14.1 |
| Structured questionnaire | 70.9 | 66.5 | 64.4 | 67.1 | 65.4 | 64.5 | 67.1 | 67.5 | 66.0 | 66.6 | 6.5 |
| *Multimodal* | | | | | | | | | | | |
| Data fusion | 73.0 | 67.7 | 66.5 | 68.6 | 63.8 | 67.9 | 67.5 | 70.0 | 61.1 | 67.3 | 11.9 |
| Feature fusion | 68.7 | 66.6 | 62.3 | 64.9 | 61.5 | 65.6 | 65.0 | 69.9 | 56.2 | 64.5 | 13.7 |
| Soft voting | 72.8 | 69.7 | 66.5 | 69.3 | 67.0 | 67.5 | 71.5 | 71.4 | 66.5 | 69.2 | 6.4 |
| Class-aware voting | 73.2 | 69.7 | 64.9 | 69.4 | 67.0 | 67.2 | 70.3 | 72.6 | 66.5 | 69.0 | 8.3 |
| Stacking | 72.7 | 68.9 | 67.9 | 69.4 | 68.3 | 66.5 | 68.8 | 71.7 | 64.9 | 68.8 | 7.8 |
| **CSMF accuracy (%)** | | | | | | | | | | | |
| *Operational comparator* | | | | | | | | | | | |
| Baseline | 86.1 | 78.5 | 86.4 | 84.4 | 84.4 | 85.2 | 81.3 | 90.5 | 83.8 | 84.5 | 12.0 |
| *Unimodal* | | | | | | | | | | | |
| Narrative text | 90.1 | 91.5 | 89.4 | 91.2 | 88.6 | 94.3 | 90.0 | 92.7 | 87.0 | 90.5 | 7.4 |
| Structured questionnaire | 92.6 | 92.3 | 87.8 | 93.2 | 92.5 | 89.4 | 94.7 | 91.3 | 89.1 | 91.4 | 6.9 |
| *Multimodal* | | | | | | | | | | | |
| Data fusion | 90.6 | 91.4 | 95.1 | 93.2 | 86.7 | 94.1 | 93.8 | 92.2 | 87.0 | 91.6 | 8.4 |
| Feature fusion | 90.2 | 91.0 | 90.4 | 90.0 | 81.2 | 91.9 | 90.9 | 93.3 | 85.9 | 89.4 | 12.2 |
| Soft voting | 91.9 | 90.6 | 91.5 | 93.3 | 89.0 | 91.5 | 92.8 | 88.5 | 87.0 | 90.7 | 6.4 |
| Class-aware voting | 92.4 | 90.9 | 90.1 | 93.1 | 90.6 | 91.1 | 93.0 | 89.0 | 87.5 | 90.9 | 5.6 |
| Stacking | 93.7 | 90.6 | 90.1 | 92.6 | 91.6 | 92.0 | 91.1 | 89.5 | 85.3 | 90.7 | 8.3 |
| **Macro F1 (%)** | | | | | | | | | | | |
| *Operational comparator* | | | | | | | | | | | |
| Baseline | 44.7 | 40.4 | 36.5 | 41.8 | 36.1 | 38.6 | 38.7 | 42.4 | 37.4 | 39.6 | 8.6 |
| *Unimodal* | | | | | | | | | | | |
| Narrative text | 55.4 | 54.4 | 42.7 | 49.5 | 45.9 | 50.7 | 50.9 | 48.7 | 36.6 | 48.3 | 18.8 |
| Structured questionnaire | 64.5 | 60.1 | 46.1 | 53.8 | 52.9 | 51.2 | 53.9 | 53.4 | 56.8 | 54.7 | 18.3 |
| *Multimodal* | | | | | | | | | | | |
| Data fusion | 61.9 | 54.2 | 52.5 | 53.4 | 49.5 | 57.8 | 52.6 | 51.4 | 44.9 | 53.1 | 17.0 |
| Feature fusion | 58.6 | 55.2 | 49.4 | 56.1 | 49.9 | 52.7 | 50.7 | 53.3 | 38.8 | 51.6 | 19.9 |
| Soft voting | 63.9 | 60.5 | 47.2 | 57.0 | 51.9 | 55.6 | 57.7 | 52.9 | 52.8 | 55.5 | 16.7 |
| Class-aware voting | 64.0 | 60.5 | 46.5 | 56.1 | 52.7 | 55.6 | 56.0 | 57.1 | 55.5 | 56.0 | 17.6 |
| Stacking | 65.2 | 56.8 | 52.3 | 54.8 | 55.4 | 53.7 | 54.9 | 55.7 | 54.5 | 55.9 | 12.9 |

### Supplementary references

[7] Manaka T, Van Zyl T, Wade AN, Kar D. Using machine learning to fuse verbal autopsy

narratives and binary features in the analysis of deaths from hyperglycaemia. arXiv preprint. 2022. arXiv:2204.12169. Available from: https://arxiv.org/abs/2204.12169.
